# A systematic analysis of IBD GWAS loci identifies most probable causal genes impacting intestinal epithelial functions

**DOI:** 10.64898/2026.07.30.26359349

**Authors:** Isabelle Hébert-Milette, Virginie Mercier, Jean Paquette, Gabrielle Boucher, Chloé Lévesque, Philippe Goyette, John D. Rioux

## Abstract

**Background:** Genome-wide association studies have identified >200 loci associated with IBD, yet the causal gene for most remains unknown. As multiple epithelial functions have been linked with susceptibility to IBD, there is a need to prioritize candidate causal genes for functional studies in this cellular context.

**Methods:** Using a standardized definition of regions implicated by index SNPs from three GWAS studies, we categorized regions as containing: (1) a known casual gene, (2) a single gene or (3) multiple genes. We then developed an *IBD Priority Score* to rank genes based on genetic, genomic and functional data. We next developed and applied an *Epithelial Priority Score*, based on expression patterns and quantitative traits, to prioritize genes for functional validation in epithelial models. Two candidate genes identified through this approach were tested for their impact on viral response pathways in HT-29 cells.

**Results:** The *IBD Priority Score* prioritized a single gene in 71 of the 104 regions containing multiple genes. The *Epithelial Priority Score* identified 31 epithelial candidates. Functional studies demonstrated that IRF6 enhanced, whereas IRF8 suppressed, antiviral responses in intestinal epithelial cells stimulated with Poly(I:C).

**Conclusions:** Combining multiple genetic, genomic, and functional data is a useful approach for prioritizing the most likely causal gene within IBD GWAS loci, and for prioritizing functional validation studies in epithelial cells and tissues. Moreover, we provide functional evidence for two IBD genes playing a role in the regulation of anti-viral responses in intestinal epithelial cells.

**Lay Summary:** Genetic studies have played an important role in identifying disease pathways. Cells lining the gut, epithelial cells, play an important role in IBD. The current study provides an approach for identifying key IBD pathways in these cells.

**Key Messages:** *What is already known?:* GWAS have identified more than 200 IBD susceptibility loci, but the causal genes remain unknown for most regions, and growing evidence implicates intestinal epithelial dysfunction in the development of IBD.

*What is new here?:* We developed an approach that prioritizes likely causal genes within IBD loci and functionally demonstrated that two of these (IRF6 and IRF8) are important for their role in antiviral pathways in human intestinal epithelial cells.

*How can this study help patient care?:* Speeding up the discovery of genes and functions relevant to IBD susceptibility pathways in intestinal epithelial cells will accelerate the discovery of novel therapies that are complementary to existing advanced therapies that target genes and functions in immune cells.

## Introduction

Inflammatory bowel diseases (IBD), which includes Crohn’s disease (CD) and ulcerative colitis (UC), are complex chronic intestinal inflammatory disorders caused by genetic and environmental factors (1). IBD involves multiple cell types, including multiple innate and adaptive immune cells, intestinal epithelial cells (IECs), and mesenchymal cells, and is associated with an altered gut microbiota. Genome-wide association studies (GWAS) have associated over 200 loci with IBD (2–4) which have led to the discovery of multiple pathways implicated in disease development. Notably, multiple immune-related functions have been linked to IBD. NOD2, a protein implicated in the recognition of pathogens that can stimulate an innate immune response, is encoded by an IBD-associated gene, *CARD15* (5). Also implicated in IBD development is the interleukin (IL)-23 axis (6). IL-23 is secreted by macrophages to induce IL-22 production by T cells (6). IL-22 then induces epithelial cells to stimulate neutrophil recruitment and inflammation (6). Multiple existing IBD treatments target the immune system and its functions. For example, thiopurines targets T cell activation and survival while Vedolizumab targets immune cell migration towards the intestine (7). Moreover, treatment that targets pro-inflammatory cytokines, like the anti-TNF-α and anti-IL-12/-23 antibody treatments have been the most widely used biologic therapies in IBD (7).

Genetic associations in IBD have also highlighted the role of the epithelial barrier in disease susceptibility. The epithelial barrier is important to limit the interactions between the immune system and the luminal microbes (8). IECs sense microbial antigens and activate immune cells accordingly (8). Moreover, Goblet cells and Paneth cells, which are specialized epithelial cells, control microbes by secreting the intestinal mucus and antimicrobial peptides, respectively. IECs are also implicated in the transport of soluble immunoglobulin A (sIGA) to the lumen to entrap microbes and promote their clearance (1). Finally, the epithelial barrier is essential for the regulation of the absorption of nutrients and water (1). In recent years, the epithelial functions of multiple IBD-associated genes have been studied (1). For example, *HNF4A* encodes a transcription factor involved in the regulation of IEC differentiation and intestinal morphogenesis (9, 10). Moreover, C1orf106 was shown to affect epithelial permeability by regulating *adherens* junction stability (11). NOD2 IBD-associated variants are associated with an increased barrier permeability (1). Moreover, NOD2 and ATG16L1 are associated with abnormal lysozyme localization and secretion (12, 13). Finally, *DUSP16* is an IBD-associated gene that regulates *PIGR* expression and therefore, the transport of polymeric IgA to the intestinal lumen (10). However, despite the emergence of IBD-associated epithelial functions, there is still no available treatment against IBD targeting IEC functions (14).

While most patients with IBD are diagnosed when they are between 18 and 35 years old (15), monogenic forms of IBD have also been identified in young patients (16). Specifically, Very Early Onset (VEO)-IBD is characterized by a severe and aggressive presentation of the disease before 6 years of age, with over 75 germline mutations known to cause VEO-IBD (17). Several genes associated with VEO-IBD have established immune-related functions, including *IL10* and the genes encoding its receptors (16, 18). IL-10 is an anti-inflammatory cytokine stimulating the expression of other anti-inflammatory genes and inhibiting the release of the pro-inflammatory cytokine TNF-α (18). Studies have also shown the role of VEO-IBD-associated genes in the maintenance of the epithelial barrier (16). Notably, TTC7A and SLC9A3 have been shown to regulate the plasma membrane composition and ion exchange at the cell membrane, respectively, and FERMT1 regulates cell adhesion to the extracellular matrix (16). Interestingly, multiple genes like *SLC9A3* and *FERMT1* are associated with both IBD and VEO-IBD (2, 4, 16), suggesting a partial overlap between the functions associated with monogenic and polygenic forms of the disease. These studies demonstrate that IEC and immune cell functions are important in both IBD and VEO-IBD development.

Despite the success of GWAS and follow-up genetic and functional studies, the causal gene within most IBD-associated loci remains to be identified (19). This challenge is not specific to IBD but rather is common to most complex traits and is due to a couple of common factors (20). First, the most associated single nucleotide polymorphism (SNP) identified in the GWAS is often not causal but is in linkage disequilibrium (LD) with the causal variant in the region (20). Finer genetic mapping, sequencing, and/or functional validation is therefore essential to identify which variant in the haplotype is the causal variant and the causal gene for the disease or phenotype (20). Second, causal variants that affect gene expression rather than protein sequence may prove to be difficult to identify if relying solely on sequence features of well-characterized regulatory elements, as this information is still incomplete and potentially tissue-dependent (20, 21). Multiple methods exist to address these challenges and to prioritize genes for future studies, but their efficacy varies (22). Combining multiple methods seems to give a better prediction success rate (22). Locus-based methods consider the distance of the gene from the GWAS SNP, the presence of an expression quantitative trait loci (eQTL) and the presence of regulatory regions (23). Network-based methods look at the protein functions and interactions of the genes present in the loci to find similar functions or pathways (24).

Rather than developing a generic approach to the problem we developed the *IBD Priority Score*, which is a straightforward approach to prioritize genes within IBD GWAS loci that takes advantage of current knowledge regarding biological pathways associated with causal IBD genes that have been functionally validated, linkage disequilibrium in the region, overlap with the genetic causes of VEO-IBD, and presence of coding variants correlated with the index GWAS SNP. With the objective of prioritizing the most likely causal gene per locus, we applied this approach to published IBD GWAS regions containing multiple genes and prioritized a single gene per locus.

As with other strategies, the prioritized candidate causal genes require functional validation. Given that gene expression and functions can be tissue/cell dependent, it is preferable that functional validation studies be performed in the most relevant cellular context possible (21). Consequently, the next step in the proposed approach is to prioritize top candidates per locus based on known tissue and single cell expression patterns and quantitative traits. In the current study, we have focused our approach on intestinal epithelial tissue and cells and created an *Epithelial Priority Score*. In doing so, we prioritized 169 genes for functional studies in epithelial tissues and/or cells. Among these, we identified two genes from independent GWAS loci (*Interferon Regulatory Factors 6 and 8; IRF6* and *IRF8*) that have previously been linked to the regulation of response to viruses in other cell types and organisms, including in immune cells (*IRF8)* (25), and in murine IEC (*IRF6)* (26). To protect against viral infection, intestinal epithelial cells must be able to (1) detect invading pathogens via PRRs, (2) release types I and III interferons (IFN), and (3) switch on the expression of Interferon Stimulated Genes (ISGs) that slow down viral replication, degrade viral RNA, and stop the spread of infections (27, 28). IRFs are a family of nine proteins that regulate IFN production and subsequent ISGs expression in a cell-type specific manner (26). As IRF6 and IRF8 have not been previously reported to be involved in the regulation of the viral response pathways in human IEC, we validated these results in human IEC models. Specifically, each of these genes was constitutively expressed in a human intestinal epithelial cell line (HT-29) and their impact on the viral response pathways was assessed. We report that overexpression of IRF6 potentiates antiviral responses at every level whereas overexpression of IRF8 impaired the pathway at the detection level.

## Materials and Methods

### Identification of candidate genes within GWAS genomic regions

The IBD associated regions from three large-scale GWAS studies (2–4) were included. Gene candidates from these GWAS regions were identified as previously reported (10). Briefly, all genes that were in linkage disequilibrium with the index SNP (r2≥0.8 using the SNAP online tool on the pilot 1K genome dataset (GRCh37/hg19)) were included in the current analysis. For regions where no genes were found in linkage disequilibrium, we included the closest gene on either side of the index SNP within a 600 000 bp region on each side of the index SNP. Genomic regions were then classified into 3 categories: **Category 1** are loci with a known IBD causal gene validated with functional studies (see **Table S1, Supplementary Data Content 1**); **Category 2** are loci with only one gene in the GWAS region by this definition or had significant fine mapping evidence in support of a single gene (29) (see **Table S2, Supplementary Data Content 1**); and **Category 3** are loci containing multiple genes (see **Table S3, Supplementary Data Content 1**).

### Closest gene analysis

For the closest gene analysis, index SNPs were searched in the dbSNP database, and the closest gene was identified by measuring the distance separating the SNP from the closest extremity of the genes within the locus. If the Index SNP was located inside a gene, this gene was the closest gene. If the SNP was in an intergenic region, the gene which was the closest to the SNP was considered the closest.

### Functions in epithelial cells and IBD/VEO-IBD-associated variant identification

A PubMed search was performed with the name of the genes and the term “epithelial cells”. Abstracts were screened for epithelial functions associated with each gene. Similarly, coding sequence variants in the IBD/VEO-IBD genes were identified by performing a PubMed search with the name of each gene and “IBD variant” or “VEO-IBD variant”. The VEO-IBD gene list was extracted from (17) and compared with the genes present in the IBD GWAS regions included in this study.

### Data extraction for eQTL analysis, single cell transcriptomic analysis and expression correlation analysis

The eQTL data for each GWAS index SNP was extracted from the Genotype-Tissue Expression (GTEx) Project on July 21, 2022 using the GTEx eQTL calculator tool accessible from the GTExPortal (see **Table S4, Supplementary Data Content 1**). The analysis was done on the Colon_Sigmoid, Colon_Transverse, and Small_Intestine_Terminal_Ileum tissue categories. If there was an eQTL with a P ≤ 0.05 in one of these tissues for one of the genes present in the Index SNP region, the gene received 1 point in the epithelial ranking.

We extracted single cell annotations from ProteinAtlas. We grouped the cell annotations into four categories: colon enterocytes, colon enteroendocrine cells, intestinal goblet cells or Paneth cells (1 point); epithelial cells (excluding colon enterocytes, colon enteroendocrine, intestinal goblet cells or Paneth cells cells; 0.66 point); multiple cell types (including epithelial cells; 0.33 points) or no predicted cell type specificity (ubiquitous or low expression; 0.33 points); others cell types (0 points).

The expression correlation score of different genes to colon enterocytes and colon enteroendocrine cells was extracted from the Tissue cell RNA of ProteinAtlas on October 25, 2022. If the correlation score was above 0.3, the genes received 1 point in the epithelial ranking.

### Gene Ontology term enrichment analysis

The ClueGO tool available in the Cytoscape application was used to perform the GO-term enrichment analysis on March 8, 2023. The 21 causal genes listed in **Table 1** were included in the analysis. Pathways that had a P <0.05, at least 2 associated genes, a minimum go-term level of 3 and a maximum go-term level of 8. An enrichment test (Right-sided hypergeometric test) was performed with a Bonferroni step down P-value correction. The GO-terms were grouped into nine groups based on the kappa scores.

**Table 1.** List of genes in Category 1 GWAS regions (i.e. loci containing known causal genes)

| Locus number | Chr | Index SNP | Associated with CD/UC/IBD based on GWAS association | Gene Symbol | Coding variant | Functional validation of variants | References (PMID) | Epithelial Priority Score |
| --- | --- | --- | --- | --- | --- | --- | --- | --- |
| 1 | 1 | rs6426833 | UC | RNF186 | RNF186(R179X); RNF186(p.Ala64Thr) | Yes | 24068945; 27381925; 27503255 | 3 |
| 2 | 1 | rs11209026 | IBD | IL23R | IL23R(R381Q), IL23R(V362I), IL23R(G149R) | Yes | 28658209; 32474165 | 2 |
| 3 | 1 | rs1801274 | IBD | FCGR2A | FCGR2A(H166R) | Yes | 28658209; 22903236 | 1 |
| 4 | 1 | rs7554511 | IBD | C1orf106 | C1orf106(p.Y333F) | Yes | 21983784; 29420262 | 1.33 |
| 5 | 2 | rs2111485 | IBD | IFIH1 | IFIH1(I923V) | Yes | 28658209; 34758847 | 2 |
| 6 | 2 | rs12994997 | CD | ATG16L1 | ATG16L1(T300A) | Yes | 28658209; 24076061; 28751470 | 1.33 |
| 7 | 3 | rs3197999 | IBD | MST1 | MST1(R703C), MST1(R651X) | Yes | 28658209; 22237417 | 1.66 |
| 8 | 5 | rs11741861 | IBD | IRGM | 20-kb deletion polymorphism | Yes | 19165925 | 1 |
| 9 | 6 | rs6927022 | UC | HLA-DRB1 | HLA-DRB1*03:01 | Yes | 31518029 | 2 |
| 10 | 8 | rs7015630 | CD | RIPK2 | N/A | N/A | 25213858 | 0.33 |
| 11 | 9 | rs10781499 | IBD, VEO-IBD | CARD9 | CARD9(S12N), CARD9(1434+1G>C) | Yes | 28658209; 31696662 | 1.33 |
| 12 | 10 | rs11010067 | IBD | CUL2 | CUL2(c.IVS17+5A>G) |  | 21983784 | 1 |
| 13 | 11 | rs11230563 | IBD | CD6 | CD6(R225W) | Yes | 28658209; 23638056 | 0 |
| 14 | 12 | rs11612508 | IBD | DUSP16 | N/A | N/A | 34758847 | 0 |
| 15 | 13 | rs3764147 | CD | LACC1 | LACC1(I254V) | Yes | 28658209; 31875558 | 1 |
| 16 | 14 | rs8005161 | IBD | GPR65 | GPR65(I231L) | Yes | 28658209; 35218908 | 1 |
| 17 | 15 | rs17293632 | IBD | SMAD3 | SMAD3(I170V) + a regulatory variant | Yes | 28658209 | 0.33 |
| 18 | 16 | rs2066847 | CD | NOD2 | NOD2(R702W), NOD2(V793M), NOD2(S431L), NOD2(N289S), NOD2(N852S), NOD2(N872S), NOD2(fs1007insC), NOD2(G908R) | Yes | 28658209; 24076061 | 0.33 |
| 19 | 19 | rs11879191 | IBD | TYK2 | TYK2(P1104A) | Yes | 28658209; 23359498 | 1.66 |
| 20 | 19 | rs516246 | CD | FUT2 | FUT2(W154X), FUT2(G258S) | Yes | 28658209; 34419617 | 3 |
| 21 | 20 | rs6017342 | UC | HNF4A | N/A | N/A | 34758847 | 2 |

N.B. Please refer to Table S1 (Supplementary Data Content 1) for complete data.

Using the same tools, the association of each candidate gene to each GO-term identified previously by the causal genes GO-term analysis above was assessed. The analysis of the association of the candidate genes with the GO-terms was performed using ClueGO with the same parameters as causal genes analysis, except that we didn’t set a minimum enrichment P-value for inclusion.

### Cell lines & Infections

HEK293T/17 (ATCC CRL-11268) cell line was used for lentivirus production and maintained in DMEM (Wisent, #319-007-CL) + 10% FBS (Sigma, #F1051) +1X Penicillin-Streptomycin (Wisent, #450-201-EL). HT-29 (ATCC HTB-38) cell line was maintained in McCoy’s 5A (Wisent, #317-010-CL) + 10% FBS + 1X Penicillin-Streptomycin + 1X Glutamax supplement (ThermoFisherScientific #35050061)

Cloning, lentiviral production and transduction, antibiotic selection and cell culture were done as in Ntunzwenimana, *et al*. (10). Briefly, the ORFs of IRF6 (IOH10956) and IRF8 (IOH42114) were recombined from pEntry donor plasmid (ThermoFisher Scientific) into our modified pLVX-EF1a-IRES-PURO/eGFP with LR reaction from the Gateway recombination cloning technology (ThermoFisher Scientific, #11791-019). Each pLVX-ORF and pLVX-empty was co-transfected into HEK293T with lentiviral packaging and envelop vectors (pMISSION VSV-G and pMISSION gagpol, Sigma, #SHP001) using TransIT-LT1 transfection reagent (Mirus, #MIR2300). Forty-eight hours after transfection, lentivirus-containing media was harvested, concentrated with Lenti-X Concentrator (Takara, #631232) and resuspended with DMEM media without serum in 1/10 of the original volume. Aliquots are kept at −80°C until used.

Proliferative HT-29 at 50% confluence in 12-well plate were transduced in triplicate for each ORFs and empty vector lentivirus with minimal infection media (McCoy5a + 1% FBS + 8 µg/ml Hexadimethrine Bromide (Sigma, #H9268)). Forty-eight hours later, puromycin (ThermoFisher Scientific, #P9620) was added to select the cells that were successfully transduced. As soon as the non-infected control cells were dead, puromycin concentration was reduced to maintain selection. For each different set of ORF transduction, three independent cell lines were generated. RNA and protein were harvested for each line to test ORFs expression by RT-qPCR and Western blot respectively. Cell lines were thereafter referred as “IRF6” or “IRF8” for HT-29 clone replicates expressing the IRF6 or IRF8 ORFs, respectively, and “Control” for the HT-29 clone replicates transfected with the empty vector.

### Treatment with Poly(I:C) and interferon-β

Clones of the colorectal adenocarcinoma cell line HT-29 stably expressing either IRF6, IRF8 or empty viral vector were transiently transfected with Poly(I:C) or treated with interferon-β (IFN-β) (Thermofisher Scientific #300-02BC-5UG) to study innate antiviral defense mechanisms as follows:

Cells (3.5 × 10^5^) were plated in 12-well plates and incubated for 24h in complete McCoy’s media containing 10% FBS, 1X penicillin-streptomycin and 1X Glutamax. Then cells were washed with D-PBS and incubated in reduced serum (1% FBS) McCoy’s media overnight. For the Poly(I:C) stimulation, cells were transfected with 5 µg High Molecular Weight Poly(I:C) VacciGrade (InVivogen, #vac-pic) using TransIT-LT1 (Mirus Bio, #MIR2300) and incubated for 4h, 8h, 12h and 24h. For the IFN-β stimulation, cells were treated with either 1 or 10 ng/mL IFN-β for 2h, 4h, 8h, and 24h. At each point, cells were lysed for RNA extraction as explained in the RNA extraction and qPCR section. For the STAT1 activation assay, cells were either transfected with 5 µg Poly(I:C) for 16 hours or treated for 40 hours with 50 ng/mL IFN-γ (Sigma-Aldrich, #SRP3058). Cells were then lysed for protein extraction as explained in the section regarding Western Blots.

### RNA extraction and qPCR

Cells were washed with D-PBS and lysed using RLT buffer from the RNA Plus mini kit (QIAGEN, #74136), supplemented with beta-mercaptoethanol. Lysates were then homogenized with a QIAshredder column (QIAGEN, #79656) and RNA was extracted according to manufacturer’s protocol with a DNase 1 (QIAGEN, #79254) treatment step between two RW1 washes. RNA was quantified by absorbance on a Take3 microvolume plate (BioTek) and 2 µg were reverse transcribed using a HighCapacity cDNA RT kit (ThermoFisher Scientific, #4368814). The cDNA was amplified by qPCR using the PowerUp SYBR Green Master mix reagent according to the manufacturer’s recommendations (ThermoFisher Scientific, #A25742) and with the QuantStudio 6 thermal Real time PCR system. Relative expression data were normalized to expression of the beta-actin gene. All the primer sequences are listed in **Table S5 (Supplementary Data Content 1**).

### Western Blots

After treatment, cells were washed and lysed in RIPA buffer containing Halt Protease and Phosphatase Inhibitor Cocktail, ThermoFisher, Scientific, #78443). Protein concentration was determined using Pierce BCA Protein Assay Kit according to manufacturer’s protocol (ThermoFisher Scientific, #23227) and the absorbance read on a Synergy 2 plate reader (Biotek). Laemmli sample buffer (Bio-Rad, #1610747), supplemented with beta-mercaptoethanol, was added to 10 ug of protein per lane and the samples were heated at 95°C for 10 min before being loaded onto a denaturing SDS-PAGE and electro-transferred onto nitrocellulose membranes. After blocking the membrane with Tris-buffered saline (TBS) supplemented with 0.1% Tween-20 (TBST) and 5% skim milk for 30 min, membranes were incubated in a diluted primary antibody O/N at 4°C. The membranes were then washed three times with TBST and incubated in the diluted HRP-labeled secondary antibody for 1 hour at room temperature. After three additional washes with TBST, proteins were revealed by chemiluminescence using the Western Blot Lightning Plus-ECL according to the manufacturer’s instructions (PerkinElmer, #NEL103001EA). Blots were analyzed with a ChemiDoc Imaging System and Image Lab Software (Bio-Rad). All the antibodies used are listed in **Table S6 (Supplementary Data Content 1**).

### Statistical analysis

#### qPCR

After normalization of each sample by the reference gene, plate effects were corrected by dividing each plate by its geometric mean. A common relative quantity scale was then obtained by dividing all samples by the average of one of the conditions, with conservation of fold-changes. Statistical analyses were performed on the logarithmic scale, so that the results on the original scale represent fold-changes. Paired design and repeated measures were taken into account in the analysis. As a first illustration of ratio vs control, differences between induced samples and the corresponding empty vector samples were computed. Mean and standard error were computed on these differences for each condition and illustrated on the original scale as geometric mean. We then performed repeated measures Anova, under the hypotheses of sphericity and normality. Marginal 95% confidence intervals were computed for the contrasts being investigated, that is, for the difference in treatment effects after gene induction, compared to empty vector. No correction was done for multiple testing on the confidence intervals.

#### pSTAT1

After normalization of each sample by their reference protein, systematic Western gel effects were corrected by dividing each gel by its median value. Each gel contained all conditions from a single replicate for STAT1 or pSTAT1. One outlier for pSTAT1 that was much lower than the other values was replaced with a small value just below the range of observed data. A common relative scale was then obtained by dividing all samples by the average of one of the conditions, with conservation of fold-changes. Ratio between pSTAT1 and STAT1 were computed for each condition and clone. Statistical analyses were performed on the logarithmic scale, so that the results on the original scale represent fold-changes. Paired design and repeated measures were taken into account in the analysis. Mean and standard error were computed for each condition and illustrated on the original scale as geometric mean.

## Results

### Definition and categorization of GWAS loci

To determine which gene should be prioritized in each IBD-associated region, we developed an IBD priority score based on genetic and functional data. We started our analysis by defining the GWAS loci based on linkage disequilibrium and physical distance surrounding the 223 index SNPs identified by three large-scale GWAS studies (see Methods for details) (2–4). Next, we sorted these loci into three categories: **Category 1** are loci with a validated IBD causal gene (N=21 loci; **Table 1 and Table S1, Supplementary Data Content 1**); **Category 2** are loci with only one gene in the GWAS region (N=98 loci; **Table 2 and Table S2, Supplementary Data Content 1**); and **Category 3** are loci containing multiple genes (N=104 loci; **Table S3, Supplementary Data Content 1**). The causal genes in the first category were selected based on genome-wide significant association (P <5 × 10^−8^) and published functional validation of a coding variant associated with IBD. For certain genes like *HNF4A*, while no IBD-associated coding variant had been identified, there was genome-wide significant association and ample functional data to link it with IBD susceptibility. These causal genes and their variants are listed in **Table 1**. While this is an evolving list of genes, we considered this a reasonable list at the initiation of the project. The genes in the second category represent excellent candidate causal genes (**Table 2**). The third category represents loci where there is insufficient genetic data to implicate a single gene, thus additional genomic data could help identify the most likely candidate causal gene.

**Table 2.**
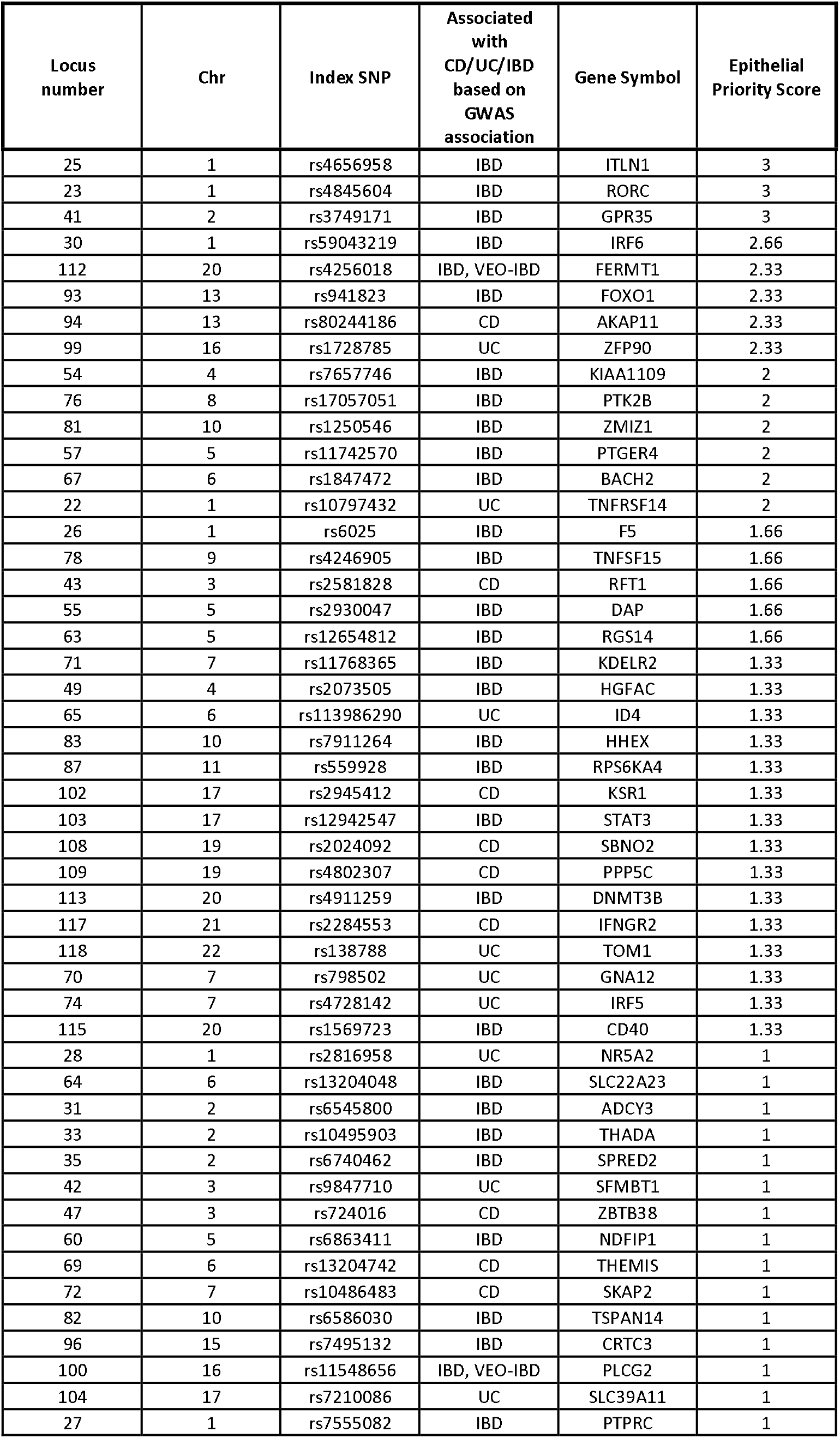
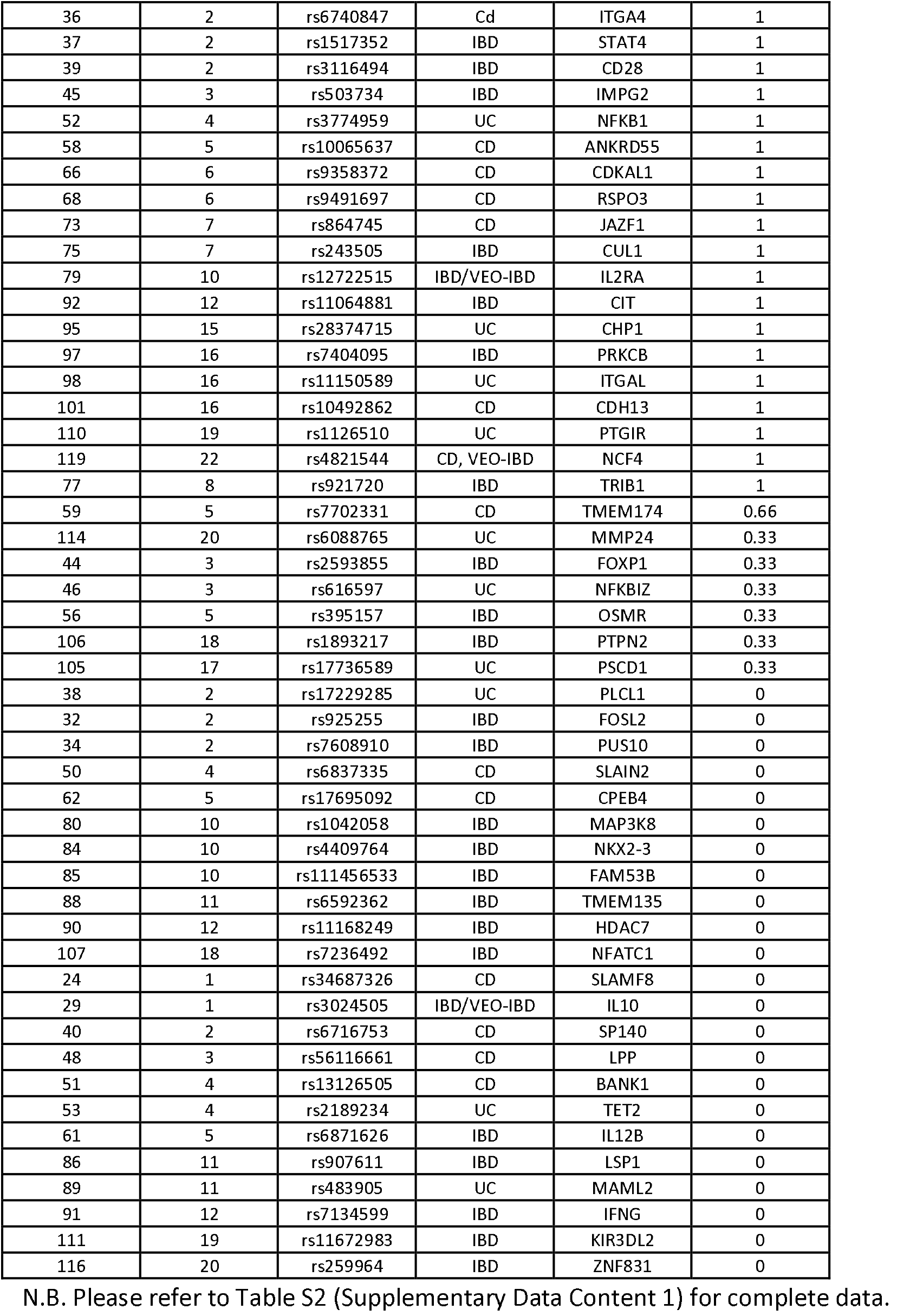
List of genes in Category 2 GWAS regions (loci containing one gene)

### Establishment of an IBD priority ranking for GWAS loci containing multiple genes

We thus ranked each gene in the third category (i.e. regions containing multiple genes) using an IBD priority score based on three criteria. The first criterion is the presence of a non-synonymous (NS) coding variant in a gene that is associated to IBD or to VEO-IBD. For the 261 genes included in the 104 regions of the third category, seven had a NS coding variant associated with IBD and two had a coding variant associated with VEO-IBD, suggesting a functional impact of the protein in IBD development. The second criterion was based on the proximity of the gene to the published GWAS index SNP (**Table S3, Supplementary Data Content 1**). Previous post-GWAS studies have provided evidence that the closest gene to the GWAS index SNP is often the causal gene of the region (30). To confirm the validity of this criterion in IBD-associated regions, we determined how many of the causal genes listed in **Table 1** are the closest gene to the IBD-associated variant in their region. We observed that 81% of the causal genes were the closest gene to the index variant in their region, supporting the use of this criterion in this study. Finally, causal genes have identified multiple biological pathways underlying IBD pathophysiology (1). The dysregulation of other proteins implicated in the same pathways could play a similar role in IBD development. Therefore, we based the third criterion on the overlap of known biological functions or pathways between the candidate genes in these regions with those identified for known causal genes. To determine the set of enriched GO-terms and functions associated with IBD, we first performed a GO-term enrichment analysis with the 21 known causal genes listed in **Table 1**. ClueGO clustered these GO-terms into nine groups of functions summarized in **Fig. S1, Supplementary Data Content 2** and in **Table S7, Supplementary Data Content 1**. We then determined which candidate genes from **Category 2** and **Category 3** identify these same GO-terms and grouped these candidate genes in the same nine functional groups (**Fig. S1, Supplementary Data Content 2 and Table S8 and Table S9, Supplementary Data Content 1**). Overall, 40% of genes that are alone in their loci and 41.1% of the genes located in the regions with multiple genes overlapped with the IBD functions identified in Category 1 genes.

For regions containing multiple genes, we summarized in **Table 3** which genes for each region should be prioritized based on our *IBD Priority Score* (see **Table S3, Supplementary Data Content 1**). This process allowed us to prioritize a single gene in 71 of the 104 regions containing multiple genes. For example, there were three regions where the maximal IBD score was obtained for a single gene: (1) *TNFAIP3*, is in a region with one other gene (*OLIG3*, IBD score of 0); (2) *TRAF3IP2*, is in a region with four other genes (all with an IBD score of 0 or 1); (3) *IL27*, is in a region with four other genes (all with an IBD score of 0 or 1) (**Table S3, Supplementary Data Content 1**). Interestingly, both TNFAIP3 and IL-27 have been studied functionally for their roles in IBD pathophysiology (31, 32). The remaining 33 regions have two or more genes with identical top prioritization scores and thus we were unable to prioritize a specific gene.

**Table 3.**
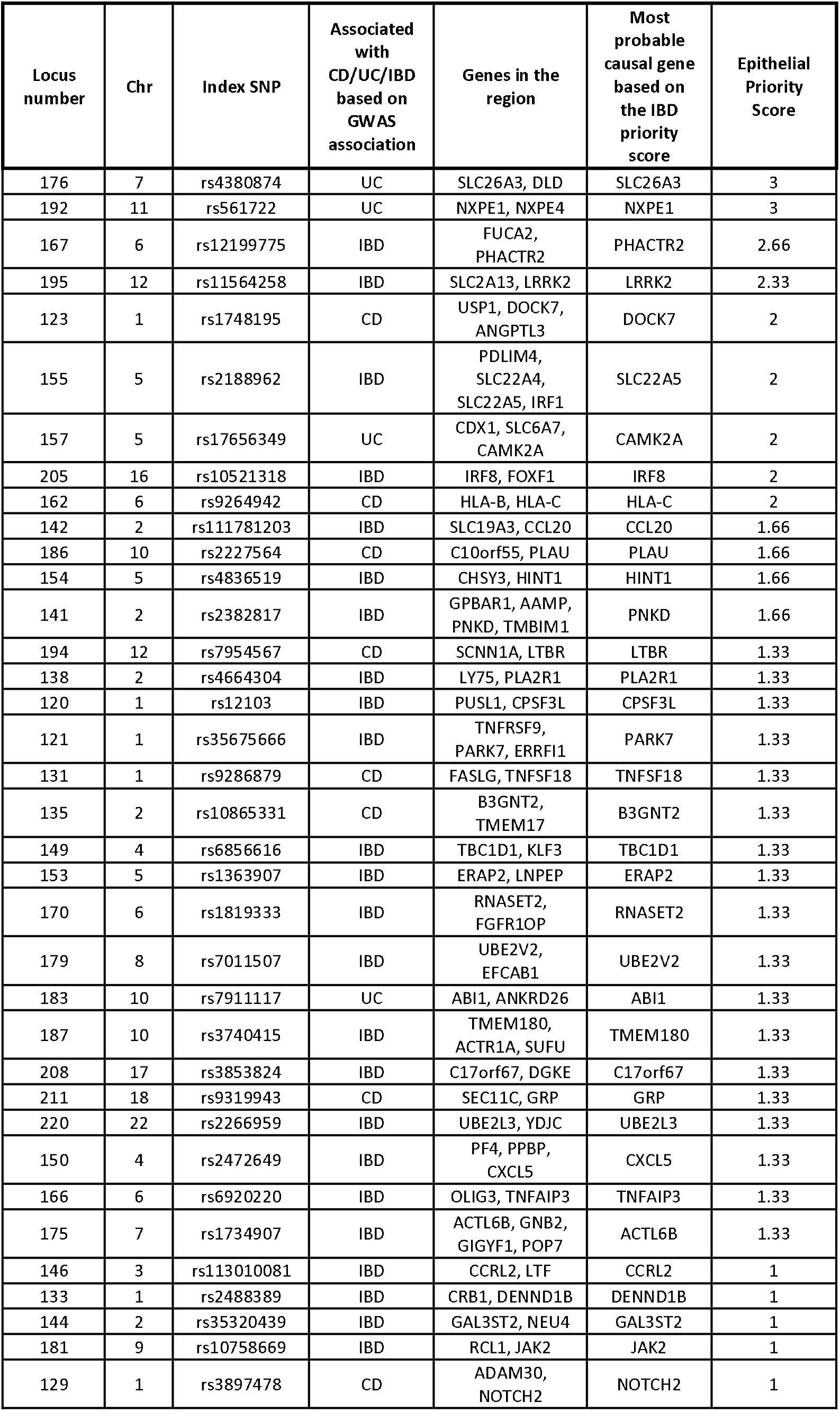
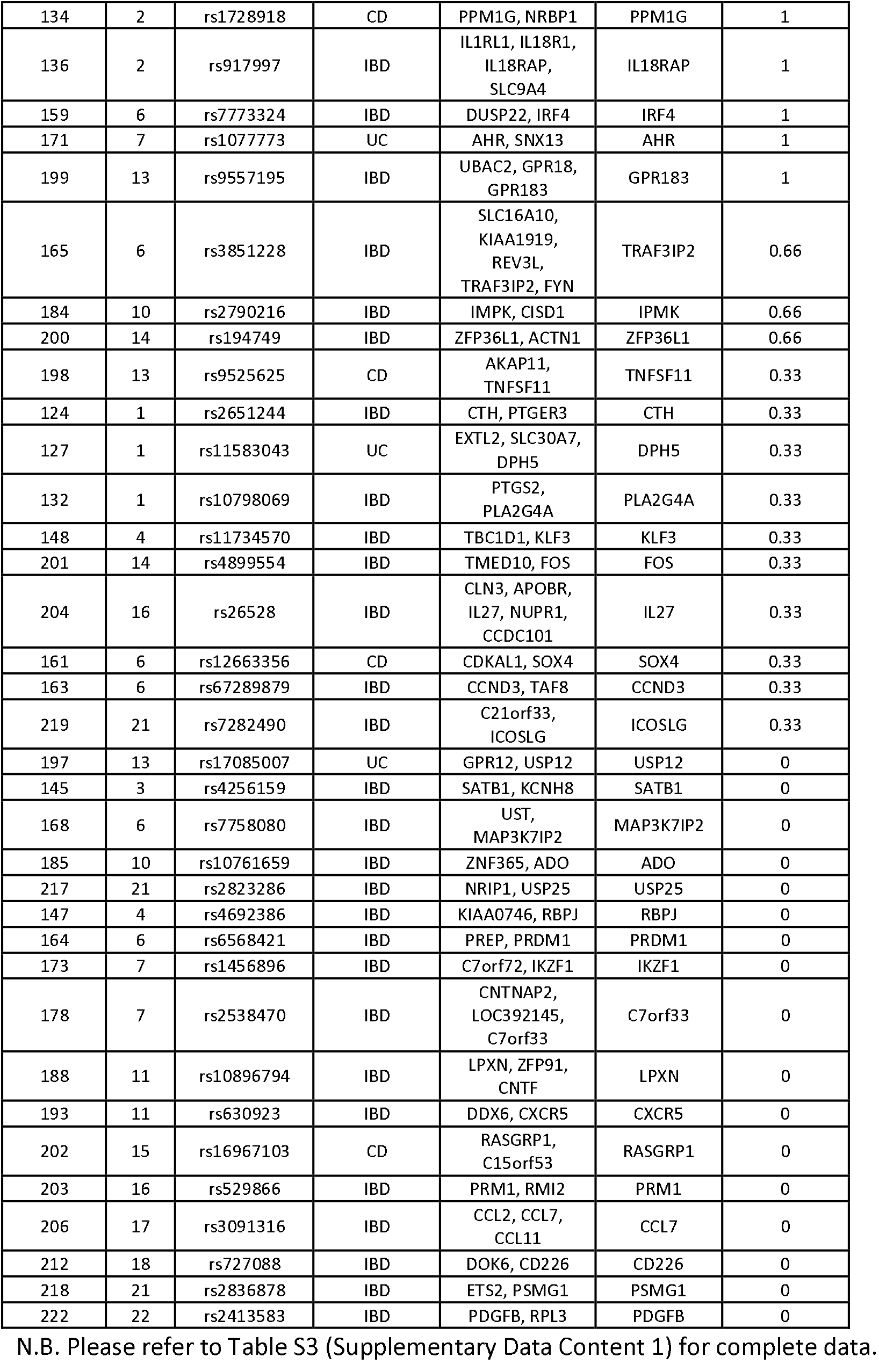
Summary of prioritized genes from Category 3 GWAS regions (ie. loci with multiple genes)

### Tissue and cell expression priority ranking

While this approach has prioritized a single gene for 71 of the 104 regions containing multiple genes, functional studies are essential to prove causality of these genes. As it is important to study the function of genes in the appropriate cellular context, we propose to use gene expression metrics to prioritize functional studies in the relevant cellular models. To illustrate this approach, we have focused on genes with epithelial expression, given the key role of epithelial cells in maintaining intestinal homeostasis and in susceptibility to IBD. As a **first** step, we determined whether the index SNP for each locus had evidence (P <0.05) in the GTEX database for being an eQTL for its corresponding candidate gene within small intestine and/or colonic tissues – this was true for 33 of the 71 prioritized genes. **Second**, we used single-cell annotations from ProteinAtlas to identify in which cell types each gene was expressed. We grouped the annotations into four categories, in decreasing order of specificity (and score weight): expressed in colon enterocytes, in colon enteroendocrine cells, in intestinal goblet cells and/or in Paneth cells (Group 1 represents 4 of the 71 prioritized genes); expression is limited to epithelial cells (Group 2, excluding genes from category 1; 8 of the 71 prioritized genes); expressed in multiple cell types (including epithelial cells) or no predicted cell type specificity (ubiquitous or low expression) (Group 3: 30 of the 71 prioritized genes); expressed in others cell types (Group 4: 29 of the 71 prioritized genes). **Third**, given the complexity of these tissues, we examined the correlation of each gene to colon enterocytes or colon enteroendocrine cells using the correlation score from ProteinAtlas generated from bulk RNA expression datasets. This correlation score was determined by the comparison of the expression profile of the genes of interest with those of reference genes for each cell type (ProteinAtlas). The higher the correlation score, the more similar the expression profile of the gene is to the expression profile of the reference genes. For our ranking, we used a correlation score cut-off of 0.3, which was sufficient to select the top ~30% of genes from category 1. Fifteen of the 71 prioritized genes passed this cut-off. The overall epithelial priority ranking for each gene is summarized in **Table S3, Supplementary Data Content 1**. The average *Epithelial Priority Score* of the 71 prioritized genes is 0.95/3, with 31 of them showing a score that is higher or equal to 1.33/3 (average for genes within Category 1) (**Table S3, Supplementary Data Content 1**), suggesting that these 31 genes might have an important role in IEC. If further prioritization is needed, the *IBD Priority Score* and the *Epithelial Priority Score* can be jointly considered within the set of 71 prioritized genes. For example, setting a minimum *IBD Priority Score* of “2” and a minimum *Epithelial Priority Score* of 1.33 identifies a list of 19 genes; the top five being *TNFAIP3, SLC22A5, CAMK2A, IRF8* and *HLA-C. TNFAIP3* encodes the A20 protein, a master regulator of NF-kappa (31). *SLC22A5* encodes OCTN2, a carnitine transporter. CAMKIIα, classically studied in the brain, is one of the four CAMKII isoforms, and has been linked to barrier functions in (33). It has been shown that IRF8 binds to interferon-stimulated response elements and regulates genes induced by type I interferon (34). *HLA-C* is a gene within the major histocompatibility complex encoding a cell-surface protein that mediates interactions with T lymphocytes and NK cells via their CD8 and KIR receptors, respectively.

Finally, we also applied the *Epithelial Priority Score* to the genes in the Category 2 loci: 34 genes had a minimum *Score* of 1.33, with four having a score over 2.66. These genes were the *intelectin-1 (ITLN1)* (Score of 3), RORC (Score of 3), GPR35 (Score of 3) and *IRF6* genes (Score of 2.66), which encode the intestinal lactoferrin receptor, a master regulator of the immune cell differentiation (Th17 in particular), a GPCR that regulates macrophage and neutrophil recruitment, and a transcription factor reported to activate the expression of the cytokine IFNβ, respectively.

### Two highly ranked genes within IBD loci regulate the response to virus pathway activation in human intestinal cells

As noted above, at least two transcription factors of the interferon regulatory factor family were prioritized as candidate causal genes having a role in intestinal epithelial cell functions: *IRF6* and *IRF8*. We set out to study the impact of IRF6 and IRF8 on their role in human epithelial cells. To do so, we genetically modified the human IEC line HT-29 to stably express elevated levels of IRF6 or IRF8 via lentiviral transduction of their respective open reading frames (ORFs).

Next, we measured the transcript and protein levels of each ORF in their respective cell lines to validate that the lentiviral transduction was successful (**Fig. S2, Supplementary Data Content 2**). We then measured the expression levels of genes involved in viral response pathways in these epithelial cell lines. As the known IBD causal gene *IFIH1* encodes the MDA5 protein which acts as an intracellular receptor for double stranded RNA (dsRNA), we have focused on this pathway (10, 29, 35). Specifically, we evaluated their impact on key pathway genes (1) **Detection:** receptors for intracellular (IFIH1) and extracellular (endosomal TLR3) dsRNA; (2) **Interferon production**: IFNb1 and IFNl1; (3) **Activation of interferon-stimulated genes (ISGs)**: to block viral entry (IFITM1), to interfere with viral protein functions by conjugating to viral and host proteins via ISGylation (ISG15), to regulate the ISG15 pathway (USP18), to inhibit viral replication and host cell survival (IFI6) and to orchestrate the innate immune response to viral infection (CCL5).

In unstimulated IECs expressing elevated levels of IRF6 or IRF8, we observed a large decrease in expression of IFITM1 and IFI6 (**Fig. S3, Supplementary Data Content 2**). In addition, in unstimulated IECs expressing elevated levels of IRF8, there was a significant decrease in TLR3 (nearly abolished), IFIH1, USP18, and ISG15. In the IRF6 expressing cells, there were pronounced decreases in the expression of IFNB1 and IFNL1.

When these cells were stimulated via transfection of Poly(I:C), used to mimic infection with a dsRNA virus, we observed a significant increase in the expression of *TLR3, IFIH1, ISG15, USP18, IFI6* and *CCL5*, in IRF6-expressing cells in comparison to control IEC at 4, 8 and 12 hours post stimulation (**Fig. 1 and S4, Supplementary Data Content 2**). The expression of IFITM1 was similar at all timepoints between IRF6-expressing and control IEC, except at 24 hours when it was lower in the former. We also observed that in IRF6-expressing cells the exposure to Poly(I:C) resulted in increased expression of *IFNB1* and *IFNL1*, however this appeared to follow a biphasic pattern with a large increase in expression at 4 hours, a substantial drop at 8 hours, followed by increases at 12 and 24 hours. The IRF8-expressing cells, on the other hand, were largely non-responsive to stimulation with Poly(I:C) at 4, 8 and 12 hours, with only a modest response at 24 hours, as assessed by the expression levels of all nine pathway marker genes. It should be noted that the expression levels of *IFNAR1* and *IL10RB*, coding for subunits of IFN type I and III receptors, respectively, were expressed at the same level between control IECs, IRF6- and IRF8-expressing cells (**Fig. S4, Supplementary Data Content 2**).

**Figure 1.**
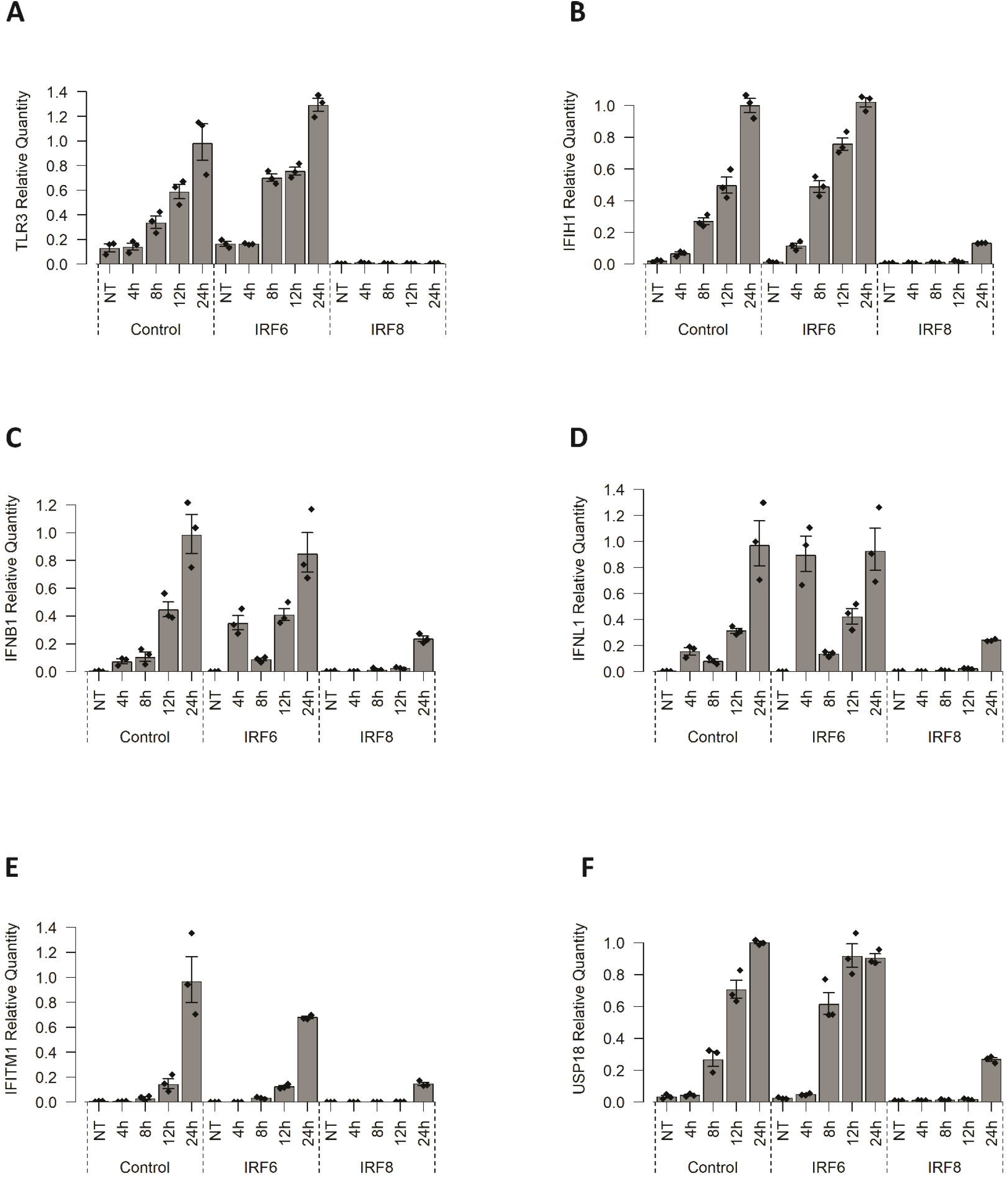
Expression level of genes involved in viral response pathways in IEC models. Control HT-29 cells, or HT-29 lines stably expressing the ORF for IRF6 or IRF8, were treated with poly (I:C) for 4 to 24 hours, NT, not treated. The expression level of key viral response pathway genes was assessed via qPCR: TLR3 (A), IFIH1 (B), IFNB1 (C), IFNL1 (D), IFITM1 (E), USP18 (F). Data from three replicates of each cell line were reported as relative expression data normalized to expression of the beta-actin gene (please see Statistical analysis in Material and Methods). The results for additional viral response pathway genes can be found in **Fig. S4, Supplementary Data Content 2.** Confidence Intervals relative to Poly (I:C) treatments for the entire set of genes can be found in **Fig. S5, Supplementary Data Content 2**. For raw data, see gene specific tables in **Supplementary Data Content 3**. Alt text: Graphical representation of the impact of IRF6 and IRF8 genes on the response of intestinal epithelial cells to Poly(I:C), a synthetic double stranded RNA. Stable expression of IRF6 resulted in an increased expression of antiviral response pathway genes TLR3, IFIH1, ISG15, USP18, IFI6 and CCL5, whereas IRF8-expressing cells, were largely non-responsive to stimulation with Poly(I:C) at 4, 8 and 12 hours, with only a modest response at 24 hours.

While the TLR3 and IFIH1 pathways have distinct adaptor protein usage and signaling cascades, they both lead to the production of interferons. Once these interferons bind to their cell surface receptors, this leads to the activation of the JAK/STAT signalling pathway. To better understand the impact of IRF6 and IRF8 on this pathway, we measured the activation of STAT1, as determined by its phosphorylation level at Tyrosine 701. We observed that the treatment of control IEC with either Poly(I:C) or IFN-γ led to an increase in phosphorylation STAT1, without noticeably changing the overall level of STAT1 protein. The increase in phosphorylation of STAT1 in response to Poly(I:C) or IFN-γ was much greater in IRF6-expressing cells (**Fig. 2**). In contrast, treatment of IRF8-expressing IEC with either Poly(I:C) or IFN-γ resulted in little-to-no increase in STAT1 activation.

**Figure 2.**
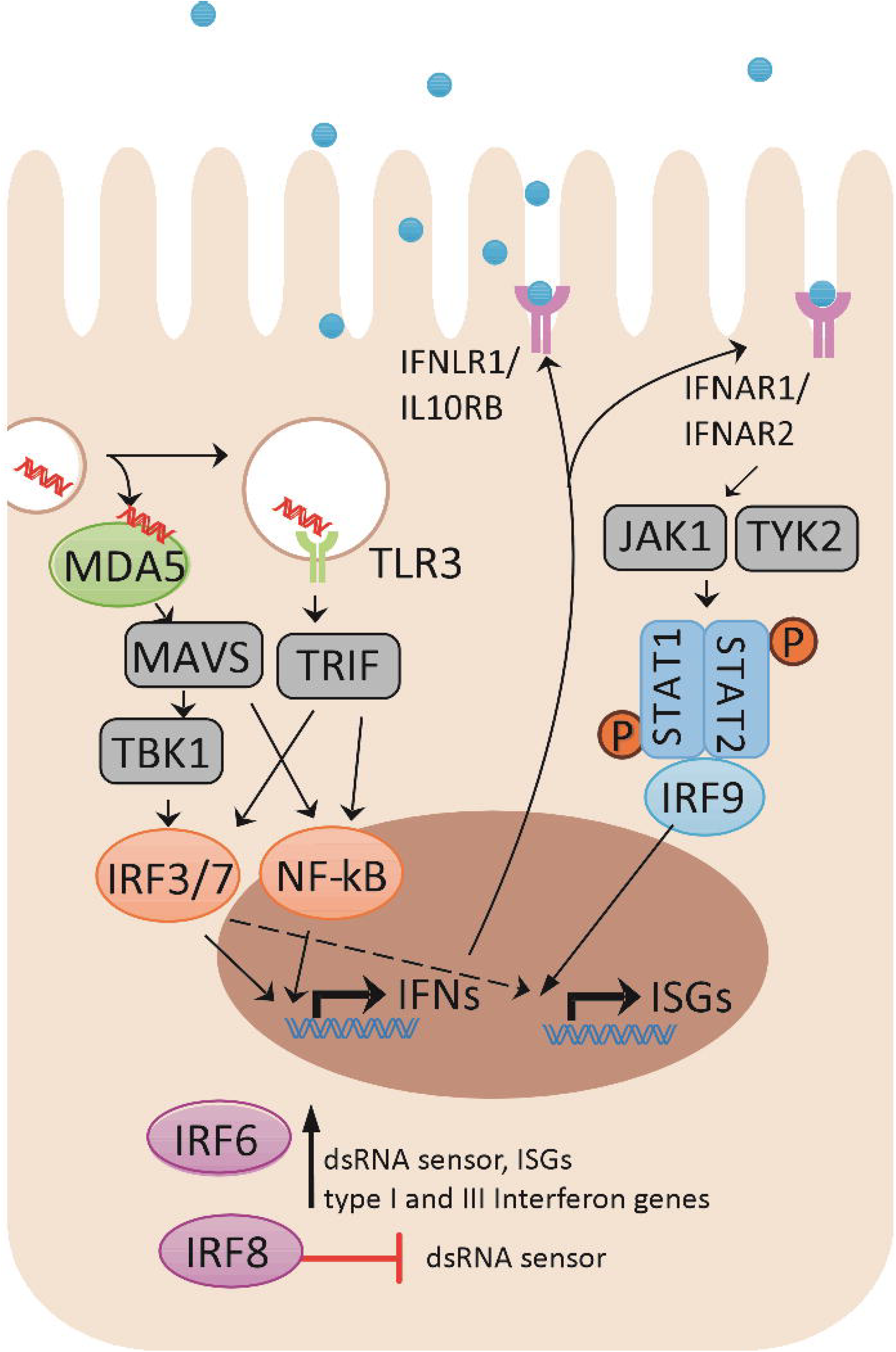
Impact of IRF6 and IRF8 on the activation of the JAK/STAT signaling pathway in IEC models. Control HT-29 cells, or HT-29 lines stably expressing the ORF for IRF6 or IRF8, were treated with poly (I:C) or IFN-γ. IFN-γ was used as a positive control since it is known that it causes the phosphorylation of STAT1 and its receptor IFNGR is broadly expressed (27, 28). Western blots were probed with anti-STAT1 **(A)**, or with anti-phospho-STAT1 **(B)** antibodies. The ratio of phosphorylated STAT1 over total STAT1 is shown in **Panel C.** For raw data see also **Supplementary Data Content 4**. Alt text: Graphical representation showing an increase in phosphorylation of STAT1 in response to Poly(I:C) or IFN-gamma was much greater in IRF6-expressing cells as compared to controls. In contrast, treatment of IRF8-expressing IEC with either Poly(I:C) or IFN-g resulted in little-to-no increase in STAT1 activation.

To further characterize the impact of IRF6 and IRF8 on anti-viral pathways in IEC, we bypassed the virus detection step by directly providing the IECs with IFNβ, a canonical type I interferon. In the control IECs, we observed that the treatment with IFNβ led to an increase in the expression of all pathway marker genes (**Fig. 3 and S6, Supplementary Data Content 2**). Similar increases in gene expression were noted in the IRF6-expressing IECs, except for the *IFNB1* and *IFNL1* genes where the increases were less pronounced. The IRF8-expressing IECs were significantly different-treatment with IFNβ led to little-to-no expression of the TLR3 receptor. These cells did express the *IFIH1* virus detection gene albeit at a lower level, thus suggesting a dramatically lower ability to respond to external viral dsRNA but still retaining a reduced capacity for detection of cytoplasmic dsRNA. In terms of interferon production in the IRF8-expressing IECs, treatment with IFNβ resulted in comparable levels of *IFNB1* expression as control cells, but reduced levels of *IFNL1*. In terms of production of ISGs, these cells expressed comparable levels of *IFITM1* as control cells, but lower levels of USP18. Of note, we detected no difference in the expression of the type-I and type-III interferon receptors *IFNAR1* and *IL10RB*, respectively, between cell lines or conditions (**Fig. S6, Supplementary Data Content 2**).

**Figure 3.**
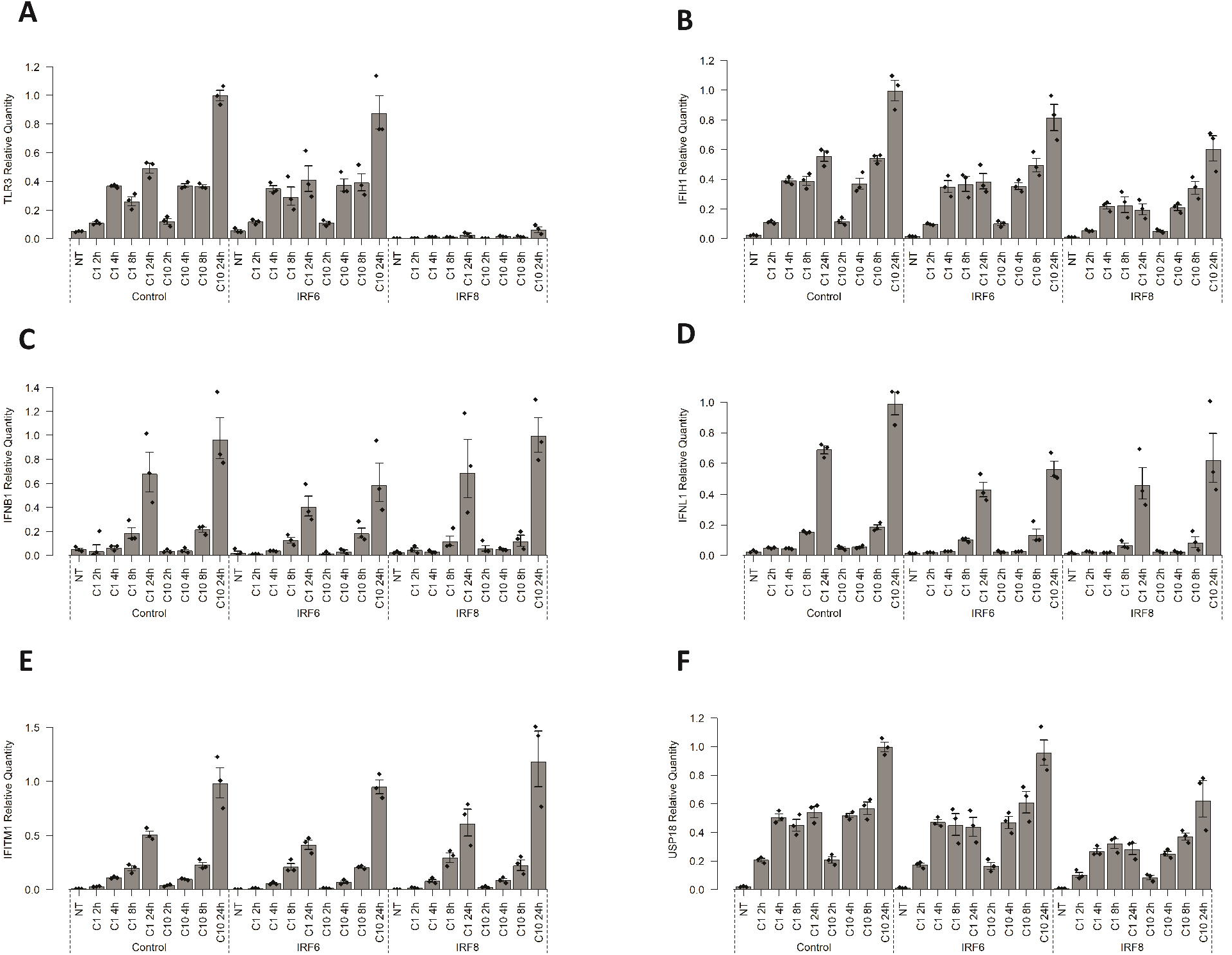
Impact of IRF6 and IRF8 on the activation of interferon responsive genes (ISGs) in IEC models. Control HT-29 cells, or HT-29 lines stably expressing the ORF for IRF6 or IRF8, were treated with IFNβ, essentially bypassing the virus detection and interferon production steps to assess the impact of IRF6 and IRF8 on ISG production. Cells were treated 2, 4, 8 and 24 hours with two different IFNβ concentrations: 1ng/ml (C1) and 10ng/ml (C10). The bar plots show expression levels determined by qPCR and reported as relative expression data normalized to expression of the beta-actin gene (please see Statistical analysis in Material and Methods): TLR3 (A), IFIH1 (B), IFNB1 (C), IFNL1 (D), IFITM1 (E), USP18 (F). Additional pathway genes are presented in **Fig. S6, Supplementary Data Content 2.** Confidence Intervals relative to IFNβ treatments for the entire set of genes can be found in **Fig. S7, Supplementary Data Content 2**. For raw data, see gene specific tables in **Supplementary Data Content 5**. Alt text: Graphical representation showing that treatment of IEC with IFNβ led to an increase in the expression of all antiviral pathway marker genes tested in IRF6-expressing IECs compared to control IECs treated with IFNβ. The IRF8-expressing IECs were significantly different-treatment with IFNβ led to little-to-no expression of the TLR3 receptor.

## Discussion

Over 200 genomic regions have been associated with IBD risk (2–4). The causal gene, however, has only been identified in a few of these regions (19). This requires determining which gene or genes are implicated by each index SNP identified by the GWAS studies, as well as functional studies to establish plausible mechanistic links with pathophysiology.

In this study, we systematically evaluated three published GWAS loci and categorized these as to containing (1) a known causal gene; (2) a single gene; (3) multiple genes. To do so, we standardized the locus definition for each index SNP identified in the GWAS studies using a combination of genetic and physical distance surrounding each index SNP. While there is no single definition for defining the genomic region surrounding each index SNP, the approach we used is in line with other studies. As much as the genes contained in Category 2 loci were immediately strong candidates as they were alone in their region, it was important to develop an approach to prioritize the genes within Category 3 loci. We thus developed an IBD priority ranking based on genetic and a functional criteria to determine which genes could be prioritized for future functional studies. The first criterion of this score is the presence of a non-synonymous (NS) coding variant in a gene that is associated to IBD or to VEO-IBD. While noncoding variants have an important role to play in IBD, the current priority score places greater emphasis on NS coding variants as these are more interpretable in the absence of other information. The relative distance from the index SNP is a powerful way of predicting the causal gene within a GWAS locus and is included as another criterion in this score (22). The closest gene is, however, not always the causal gene (22). The IBD priority score thus also includes an element that takes into account molecular and functional overlap with known IBD genes, with the premise that multiple different IBD risk genes exert their impact on a smaller set of biological pathways (e.g. innate response to microbes, cytokine signaling, etc.). To do so, we performed an enrichment analysis of the Gene Ontology (GO) terms associated with the Category 1 gene (i.e. known causal) list and determined the overlap between these and the GO terms for genes in Category 3 loci. These three elements were then combined into a single score that was used to prioritize a single gene within each locus. As such, the prioritized gene across the different Category 3 regions can have different priority scores, thus it is possible to prioritize further which loci to pursue functionally if limited at this step (e.g. throughput of functional assay, resources, etc.).

As the objective of the *IBD Priority Score* was to help select genes for functional follow-up, and that it is often important to study genes within the appropriate biological context, we next developed an approach to prioritize genes based on different expression-based criteria. Given that most current treatments for IBD focus on immune cells, and that there is an emerging interest in the potential for treatments that target the intestinal epithelium, we developed an *Epithelial Priority Score*, although this same approach can be modified for other cells or tissues of interest. This score integrates information regarding links between the GWAS index SNPs and control of gene expression in epithelial tissues (eQTLs) as well as complementary expression data from bulk and single cell RNA sequencing experiments. Approximately 40% of the genes in the Category 1-3 loci had an *IBD Priority Score* of 1.33 or greater, suggesting that many IBD genes are likely to play a role in intestinal epithelial functions. We were interested in the observation that two interferon responsive factor genes, *IRF6* and *IRF8*, were identified with the current approach as being candidate IBD genes. IRF6 has been extensively studied for its role in keratinocyte differentiation (36). The knockout of *IRF6* has also been shown to repress cell growth and cell differentiation in intestinal epithelial cell organoids from mouse (26). Moreover, that study also showed that *IRF6* KO organoids had 20 ISG upregulated and a decreased survival after type I and III IFNs treatment compared to controls, suggesting a role for IRF6 in the response to IFN pathways (26). IRF8 functions have been studied in immune cells, where it is involved in immune cell differentiation and antiviral responses (25). However, its functions in epithelial cells remained poorly understood.

We therefore studied both the IRF6 and IRF8 genes by expressing their corresponding ORFs in an established human intestinal epithelial model (HT-29) and examined the impact of the increased expression of these genes on known markers of the anti-viral response pathway. We observed that elevated expression of IRF6 or IRF8 impacted the expression of multiple genes in this pathway. In particular, in IRF8-expressing IECs, there was significantly lower expression of both receptors for dsRNA, with TLR3’s expression being abrogated completely. Not surprisingly, these IEC were nonresponsive to stimulation with Poly (I:C), apart from a modest expression of some pathway genes at 24 hours post stimulation. Moreover, we demonstrated a decreased activation of STAT1 after a Poly(I:C) treatment, supporting a repression of the pathway in these cells.

In IRF6-expressing IEC, Poly (I:C) stimulation led to a more rapid increase in expression of the dsRNA receptors TLR3 and IFIH1, as well as the ISGs USP18, ISG15, IFI6 and CCL5. The interferon genes tested in these IRF6 IECs, IFNB1 and IFNL1, also had a rapid increase that followed a biphasic pattern peaking at 4 and 24 hours post stimulation. This pattern has also been observed in innate immune cells post viral infection and corresponds to a coordinated regulatory gene network that enables an inflammatory response followed by a transition to an antiviral and tissue repair mode (37). Taken together, these results suggest that genetic variation in the IRF6 and IRF8 gene regions have a role to play in susceptibility to IBD, likely affecting how these two regulate the balance between pro-inflammatory and anti-viral & tissue repair responses in IECs (**Fig. 4**). More broadly, this is consistent with IBD susceptibility genes having an important role in anti-microbial pathways in IECs. Previously, a functional screen of genes within IBD-associated regions found evidence that *SBNO2, NFKB1*, and *IFIH1* could regulate the expression of multiple genes involved in the type 1 interferon response pathway to viral components in IEC (10). Interestingly, the impact of three IBD-associated NS coding variants in *IFIH1* negatively impacted the expression levels of multiple genes involved in the type 1 interferon response pathway (10). Moreover, in iPSC-derived human IEC carrying IBD-associated variants of *IFIH1*, the preservation of the epithelial barrier integrity was impaired after an exposure to enteric viruses compared to wild-type cells (35), suggesting that an improper regulation of the response to virus pathway could lead to greater inflammation.

**Figure 4.**
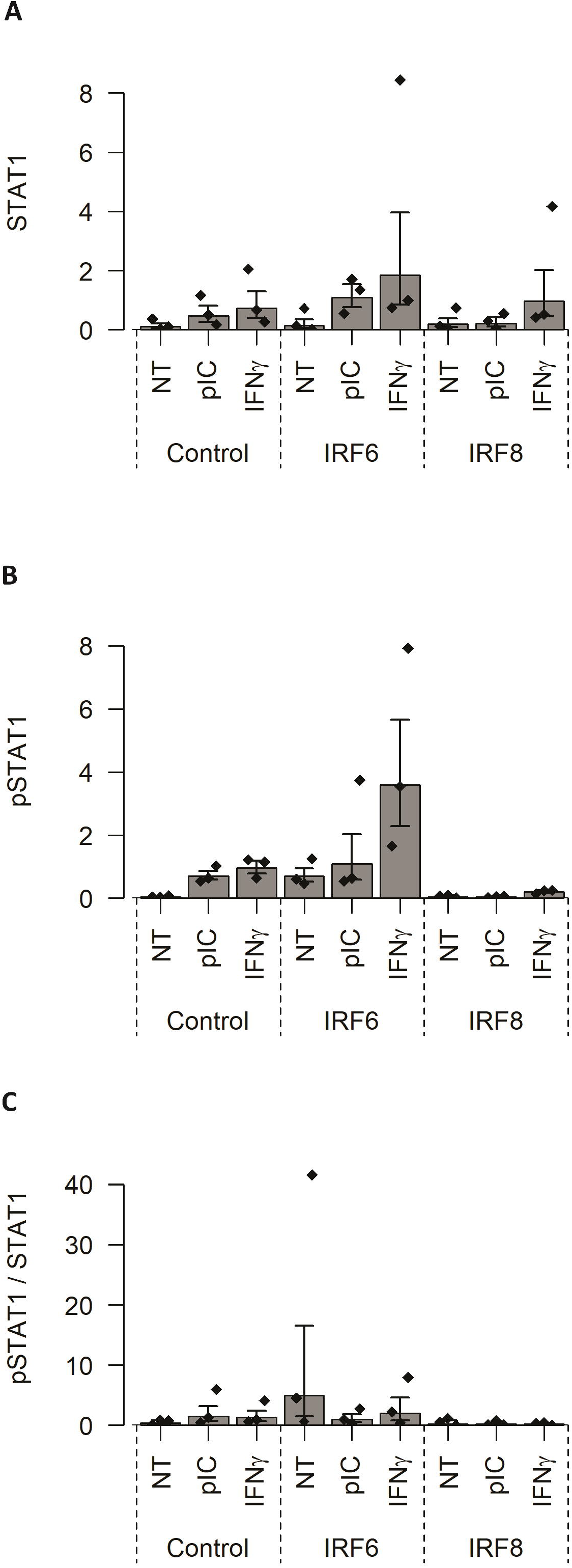
Potential role of IRF6 and IRF8 genes in susceptibility to IBD, by regulating the balance between pro-inflammatory and anti-viral & tissue repair responses in IECs. A schematic representation of the proposed impacts of IRF6 and IRF8 on the anti-viral response of IECs. Upon viral infection by dsRNA virus in IECs (or recognition of analogs such as Poly(I:C)), the receptors for this viral nucleic acid known as TLR3 (extracellular/endosomal) and MDA5 (intracellular, encoded by *IFIH1*) activate the production of types I (IFNα/β) and III (IFNλ) IFN via transcription factors like IRF3/7. Type I IFN receptors (IFNAR1/IFNAR2) and type III IFN receptors (IFNLR1/IL-10RB), activated via autocrine and paracrine mechanisms, in turn cause the formation of a complex comprised of IRF9 and phosphorylated STAT1 and STAT2. This complex then binds to IFN-stimulated response element sequences in the DNA, leading to transcription of IFN-stimulated genes (ISG). The results suggest that IRF6 impacts at the level of virus detection, IFN production and ISG expression while IRF8 would act mainly by blocking the expression of the viral dsRNA receptors TLR3 and MDA5. Alt text: A schematic representation of the proposed impacts of IRF6 and IRF8 on the anti-viral response of IECs. The results suggest that IRF6 impacts at the level of virus detection, type I and type III IFN production and ISG expression while IRF8 would act mainly by blocking the expression of the viral dsRNA receptors TLR3 and MDA5.

In summary, the approach described herein takes advantage of published knowledge regarding the genetics, genomics and functional annotations to prioritize genes within IBD GWAS loci for functional studies. We have further provided experimental evidence in support of two genes, prioritized with this approach, as being implicated in IBD pathophysiology, as well as being part of a broader set of IBD genes involved in the anti-microbial response of IECs. This can provide some confidence that this prioritization approach can successfully identify the most likely causal gene within a GWAS locus. As with any other analogous approaches, there will be false positives and false negatives, the levels of which are difficult to assess at this moment. Importantly, this approach is meant to be complementary to others and not meant to replace *in vitro* or *in vivo* approaches to screen large numbers of variants and/or validate in more complex physiological systems (38, 39). While validation of the remaining candidates will require additional studies, hopefully the lists of candidate causal genes provided herein will be useful for other researchers to apply their functional tools and strategies, as well as an approach that can be applied to future GWAS loci. Finally, we believe that this work contributes to the growing evidence of an important role for IECs in susceptibility to IBD.

## Supporting information

Supplementary Data Content 1

Supplementary Data Content 2

Supplementary Data Content 3

Supplementary Data Content 4

Supplementary Data Content 5

## Data Availability

All data produced in the present work are contained in the manuscript

## ACKNOWLEDGEMENTS

We would like to thank Julie Thompson Legault for her administrative assistance.

## ETHICAL CONSIDERATIONS

Exempt; no animal or human subjects were included in the study.

## ABBREVIATIONS

CD: Crohn’s disease
eQTL: expression quantitative trait locus
GWAS: genome-wide association studies
IBD: inflammatory bowel diseases
IEC: intestinal epithelial cells
IFN: interferons
IL: interleukins
IRF: interferon regulatory factors
ISG: interferon-stimulated genes
ORF: open reading frame
PRR: pattern recognition receptors
SNP: single nucleotide polymorphism
TLR: toll-like receptors
UC: ulcerative colitis
VEO-IBD: very early onset inflammatory bowel disease

