## Supplementary Data Content 2 for "A systematic analysis of IBD GWAS loci identifies most probable causal genes impacting intestinal epithelial functions"

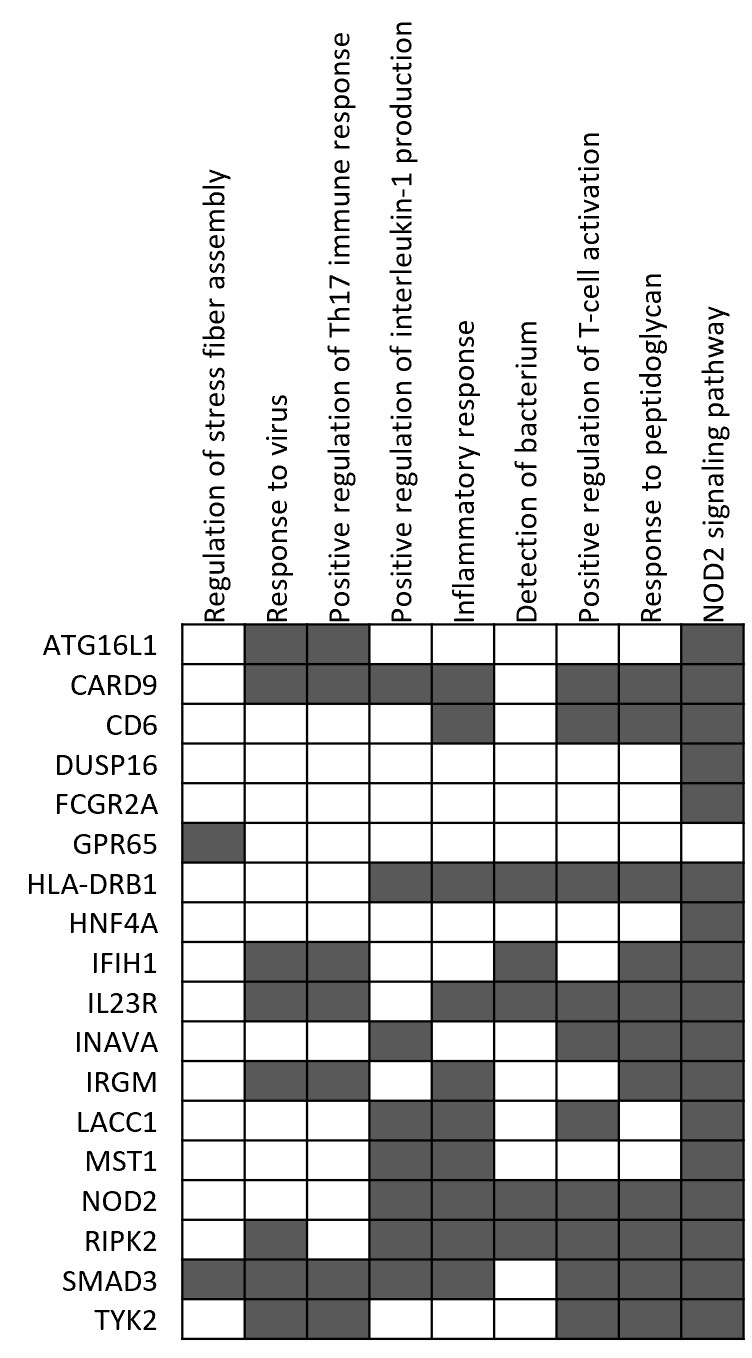

Figure S1. Panel A

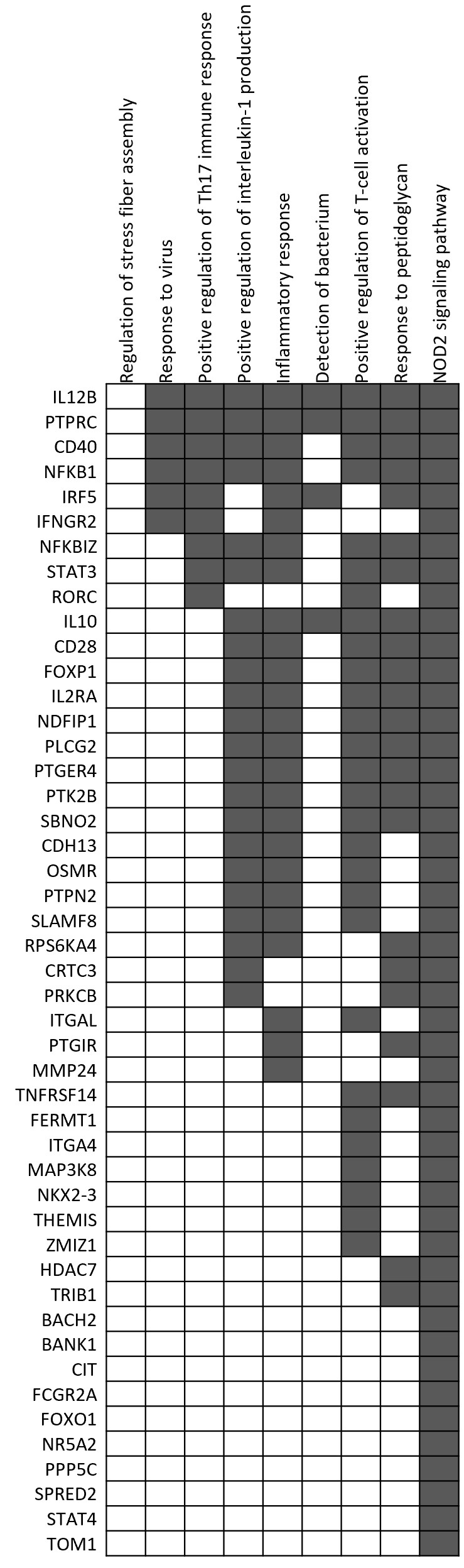

Figure S1. Panel B

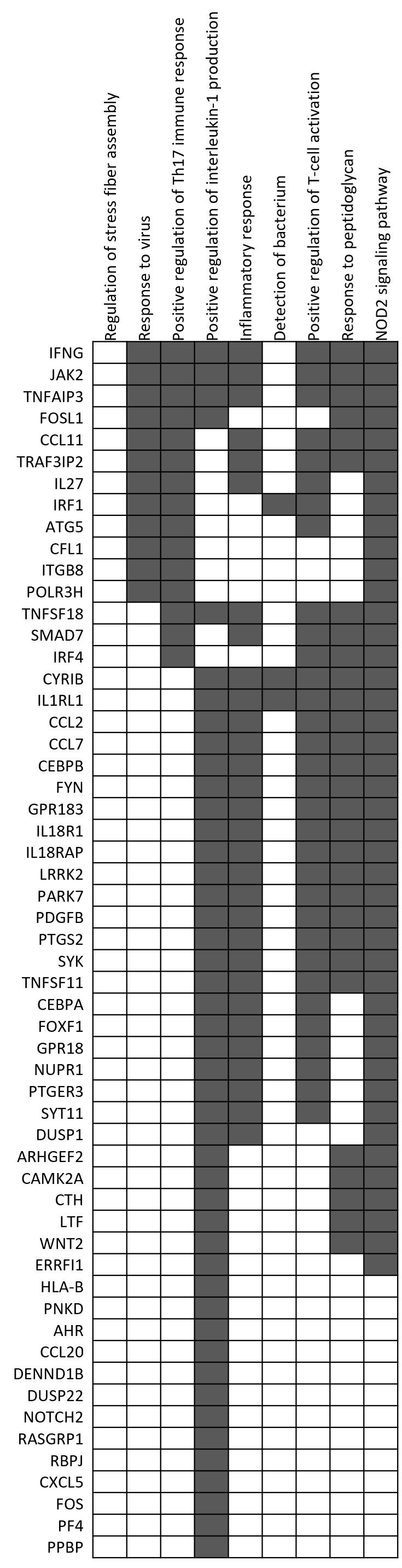

Figure S1. Panel C

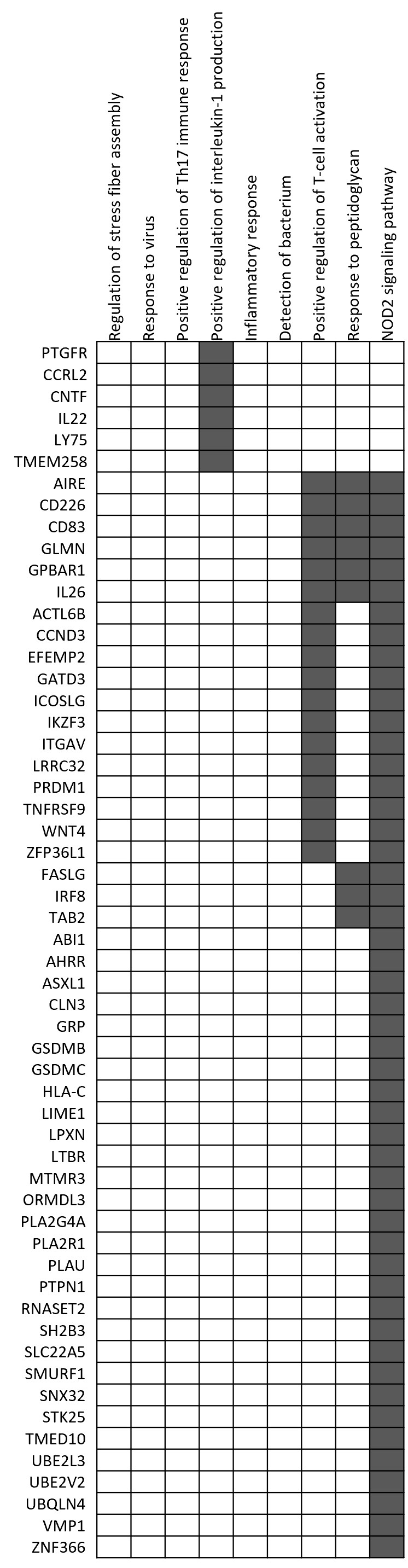

Figure S1. Panel C (Continued)

**Figure S1. Gene ontology (GO) terms associated with genes within IBD GWAS loci.** Panel A presents a heatmap of the set of enriched GO-terms, clustered in nine functional groups for the 21 known causal genes (ie Category 1) listed in Table 1. GO-term enrichment analyses were then performed on genes within Category 2 (B) and Category 3 (C) regions, organized in the same nine functional groups. Grey boxes represent presence of an enrichment of GO terms within a functional group for a given gene, thus illustrating the extent to which they overlap with functions for known causal genes. The results from these analyses can be found in **Table S2**, **S3, S7 and S8, Supplementary Data Content 1**.

Alt text: This is a graphical representation of gene ontology enrichment analyses that identified multiple terms associated with known causal IBD genes, that could be clustered into nine functional groups: Regulation of stress fiber assembly; response to virus; positive regulation of Th17 immune responses; positive regulation of interleukin-1 production; inflammatory response; detection of bacteria; positive regulation of T-cell activation; response to peptidoglycan; NOD2 signaling pathway. Two separate panels illustrate the extent to which the results from the gene ontology enrichment analyses of genes within Category 2 and Category 3 regions overlap with those.

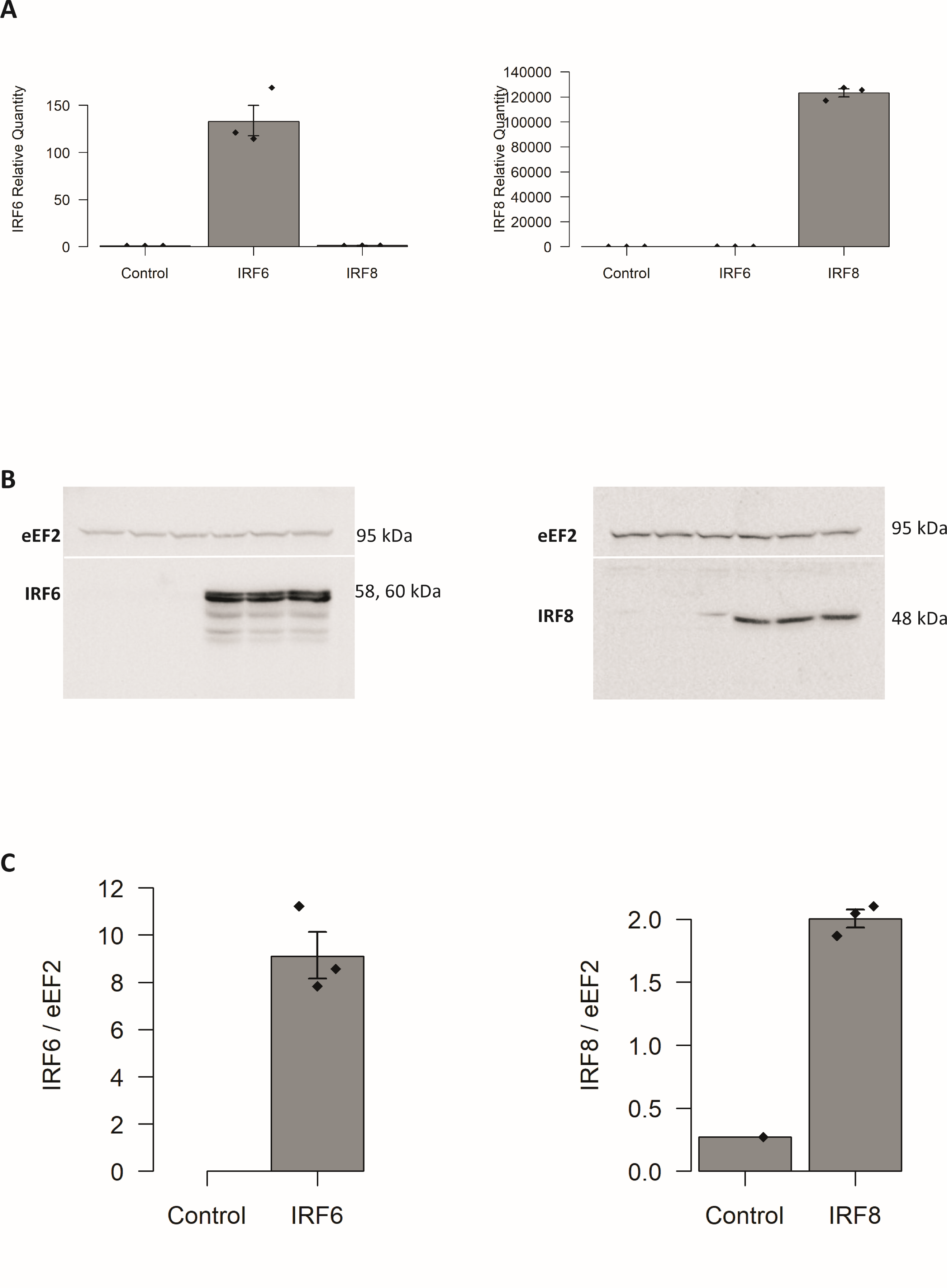

**Figure S2. Validation of the IEC models expressing IRF6 or IRF8.** Expression of transduced IRF6 and IRF8 in HT-29 cells was assessed at the RNA and protein levels as compared to control HT-29 cells. As described in Methods (Cell line & Infections), three independent replicates of HT-29 cell line were obtained. Transcript and protein levels for IRF6 and IRF8 were assessed via qPCR (**A**) and via Western Blot (**B**), respectively. Protein levels were quantified and are shown relative to eEF2 (**C**).

Alt text: This figure demonstrates that the viral transduction of the open reading frame for the IRF6 and IRF8 genes led to a significant increase of these genes, at the level of RNA and protein, in their respective cell lines.

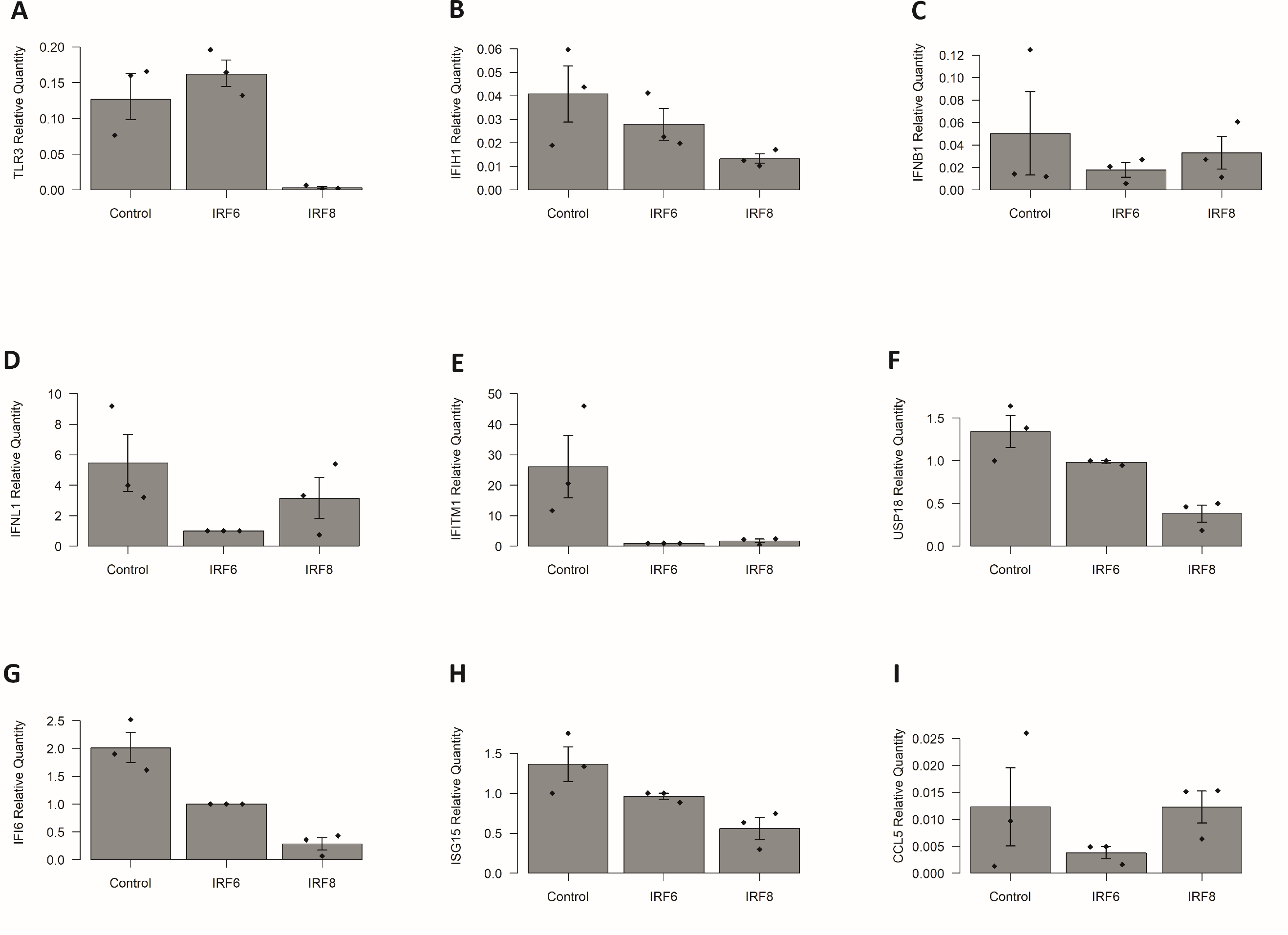

**Figure S3. Basal expression levels of antiviral response pathway genes in different IEC models.** The levels of antiviral response pathway genes were assessed in untreated control HT-29 cells, or HT-29 lines stably expressing the ORF for IRF6 or IRF8. Data from three replicates of each cell line were reported as a relative expression normalized to expression of the beta-actin gene. The data reported here were extracted from those in **Fig. 1** **and Fig. S4**. For raw data, see gene specific tables in **Supplementary Data Content 3**.

Alt text: This is a schematic representation of basal expression levels of antiviral response pathway genes in different IEC models in their unstimulated state (i.e. not stimulated with poly (I:C) or interferons). The stable expression of IRF8 resulted in decreased expression of most of the genes tested, most dramatically for TLR3, IFIH1, and IFITM1. The stable expression of IRF6, on the other hand, resulted in similar patterns of expression as the control cells, except for the lower levels of IFNL1, IFITM1 and IFI6.

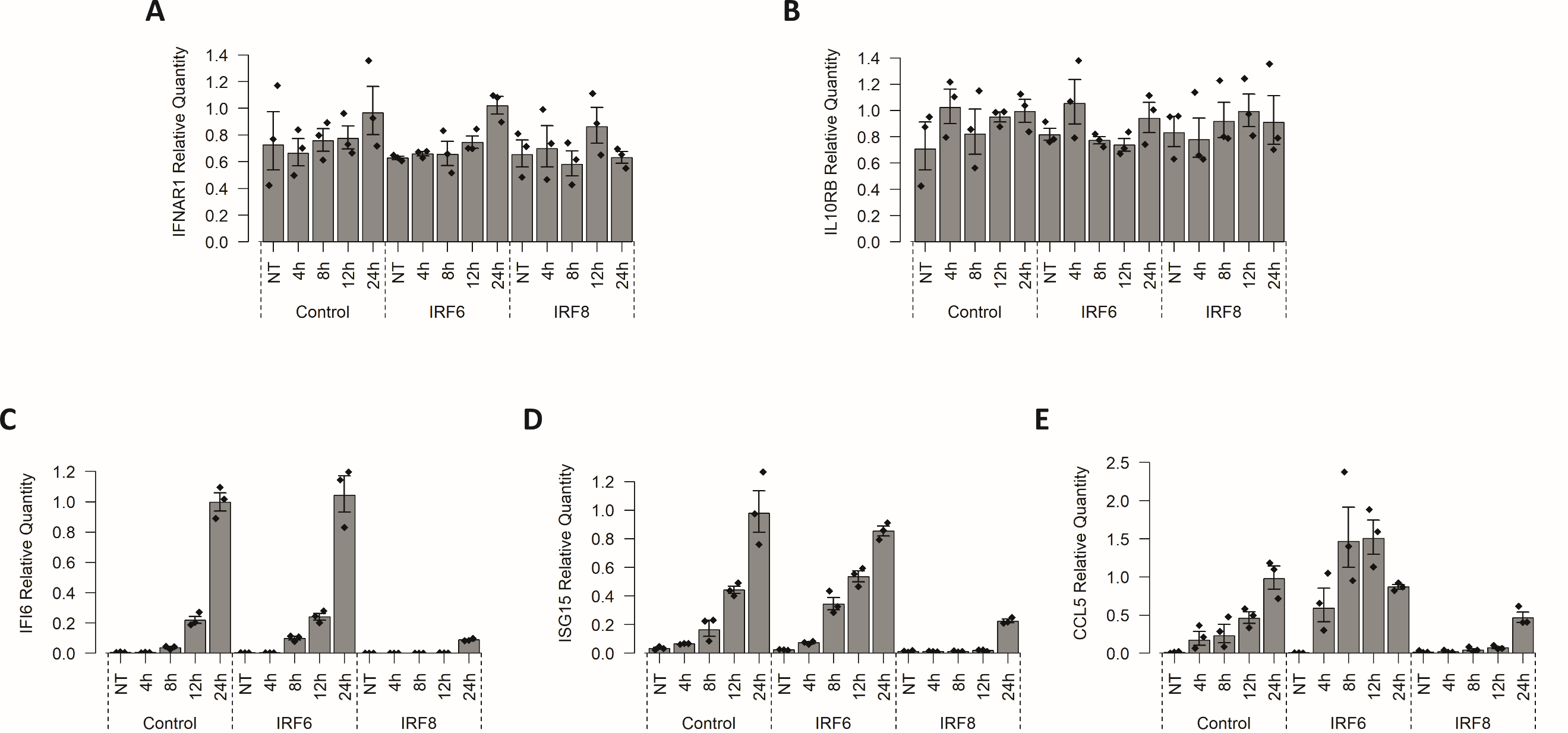

**Figure S4. Expression level of genes involved in viral response pathways in IEC lines (Additional set of pathway marker genes).** Poly (I:C) treatment as reported in **Fig. 1** and effects on additional genes: IFNAR1 (A), IL10RB (B), IFI6 (C), ISG15 (D), CCL5 (E). Confidence Intervals relative to Poly (I:C) treatments for the entire set of genes can be found in **Fig. S5**. For raw data, see gene specific tables in **Supplementary Data Content 3**.

Alt text: Graphical representation of the impact of IRF6 and IRF8 genes on the response of intestinal epithelial cells to Poly(I:C), a synthetic double stranded RNA. This is for an additional set of five genes not included in Figure 3. This figure provides evidence that stable expression of IRF6 resulted in earlier expression of the IFI6, ISG15 and CCL5 genes in response to poly (I:C) as compared to controls but had no impact on the receptors for type I or type III interferons. Stable expression of IRF8 also had no impact on the expression of the receptors for type I or type III interferons, but dramatically decreased the expression of the ISGs, namely IFI6, ISG15 and CCL5.

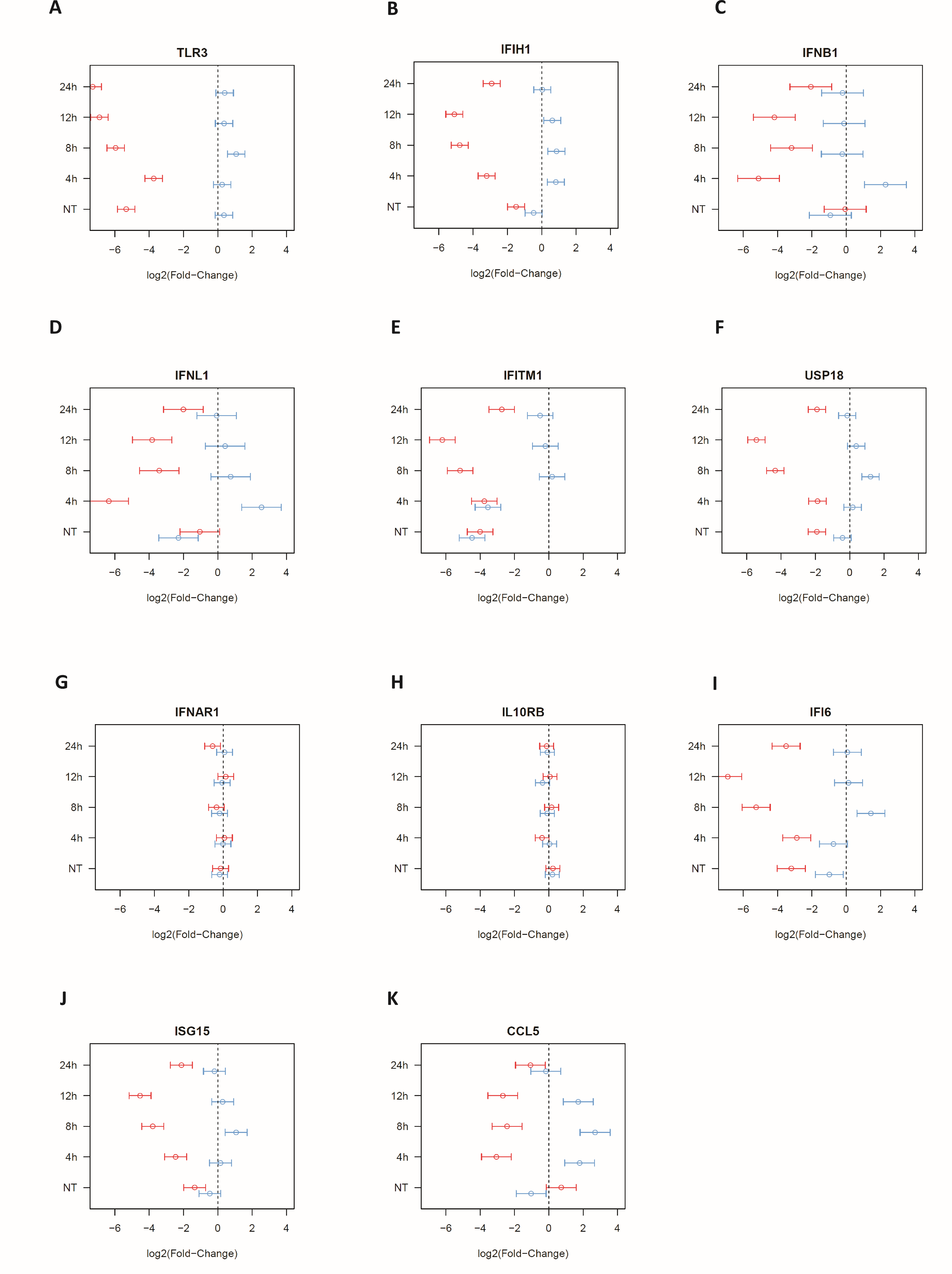

**Figure S5. Comparison of the impact of IRF6 and IRF8 on the expression level of genes involved in viral response pathways in IEC lines.** Confidence Intervals relative to Poly (I:C) treatments for the results shown in **Fig. 1 and Fig. S4**. Confidence intervals for IRF6 and IRF8 are represented by blue and red bars, respectively. For raw data, see gene specific tables in **Supplementary Data Content 3**.

Alt text: This is a graphical representation of a comparison of the effects of IRF6 and IRF8, highlighting their differential impacts on antiviral response pathways, with the former primarily increasing and /or speeding up the response, whereas IRF8 primarily depresses the response.

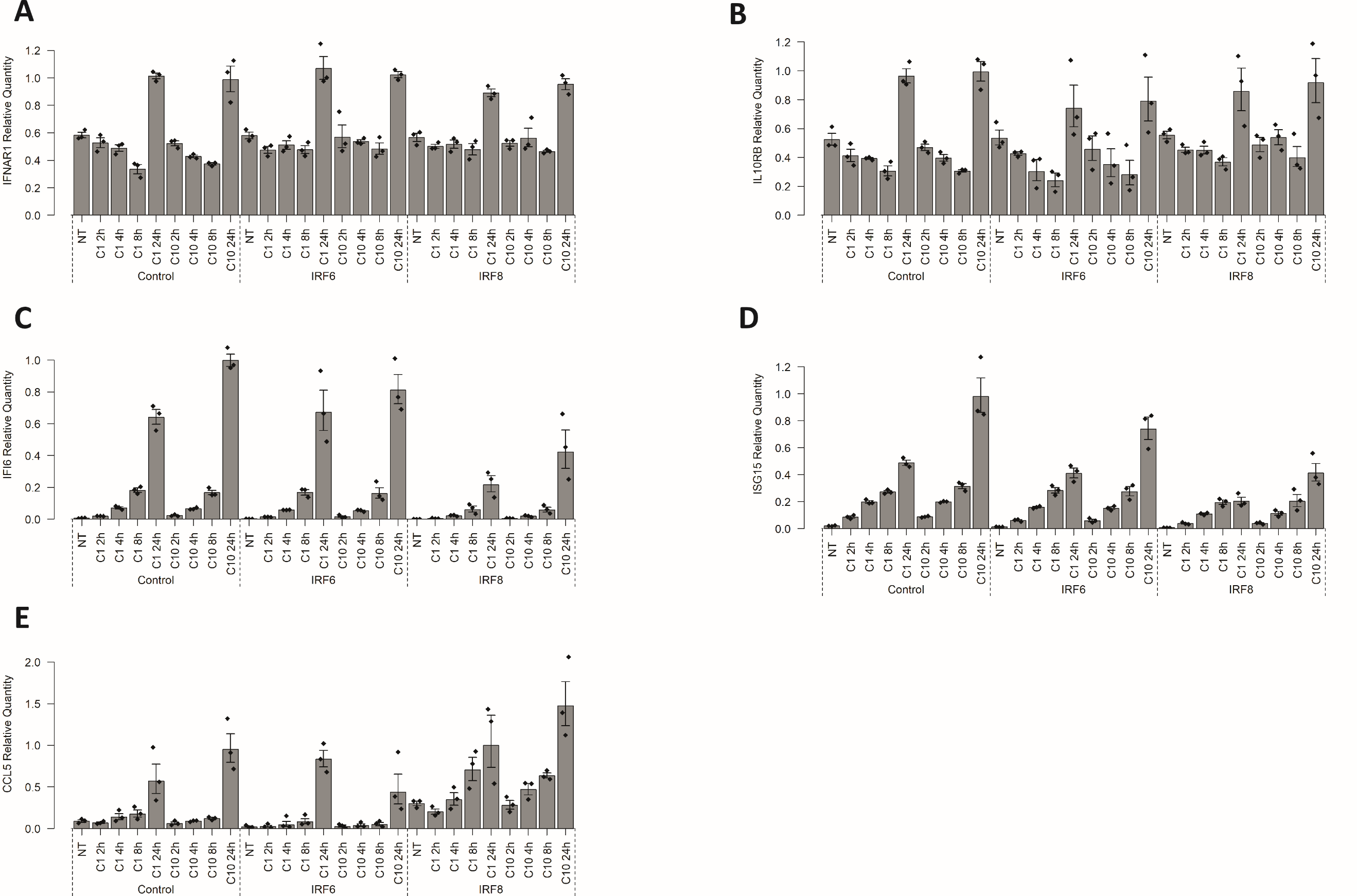

**Figure S6. Impact of IRF6 and IRF8 on the activation of interferon responsive genes (ISGs) in IEC models.** Control HT-29 cells, or HT-29 lines stably expressing the ORF for IRF6 or IRF8, were treated with IFNβ, essentially bypassing the virus detection and interferon production steps to assess the impact of IRF6 and IRF8 on ISG production. Cells were treated 2, 4, 8 and 24 hours with two different IFNβ concentrations: 1ng/ml (C1) and 10ng/ml (C10). The bar plots show expression levels determined by qPCR and reported as relative expression data normalized to expression of the beta-actin gene (please see Statistical analysis in Material and Methods): IFNAR1 (A), IL-10RB (B), IFI6 (C), ISG15 (D), CCL5 (E). These genes are in addition to those presented in **Fig. 3**. For raw data, see gene specific tables in **Supplementary Data Content 5**.

Alt text: This is an extension of Figure 3, simply a different set of genes. There was no discernible difference between cell lines expressing IRF6, IRF8 or control cell lines in terms of the expression of type I and type III interferon receptors in response to two different IFNβ concentrations. Stable expression of IRF8 decreased the responsiveness of cells to IFNβ, as assessed by the levels of ISGs IFI6 and ISG15.

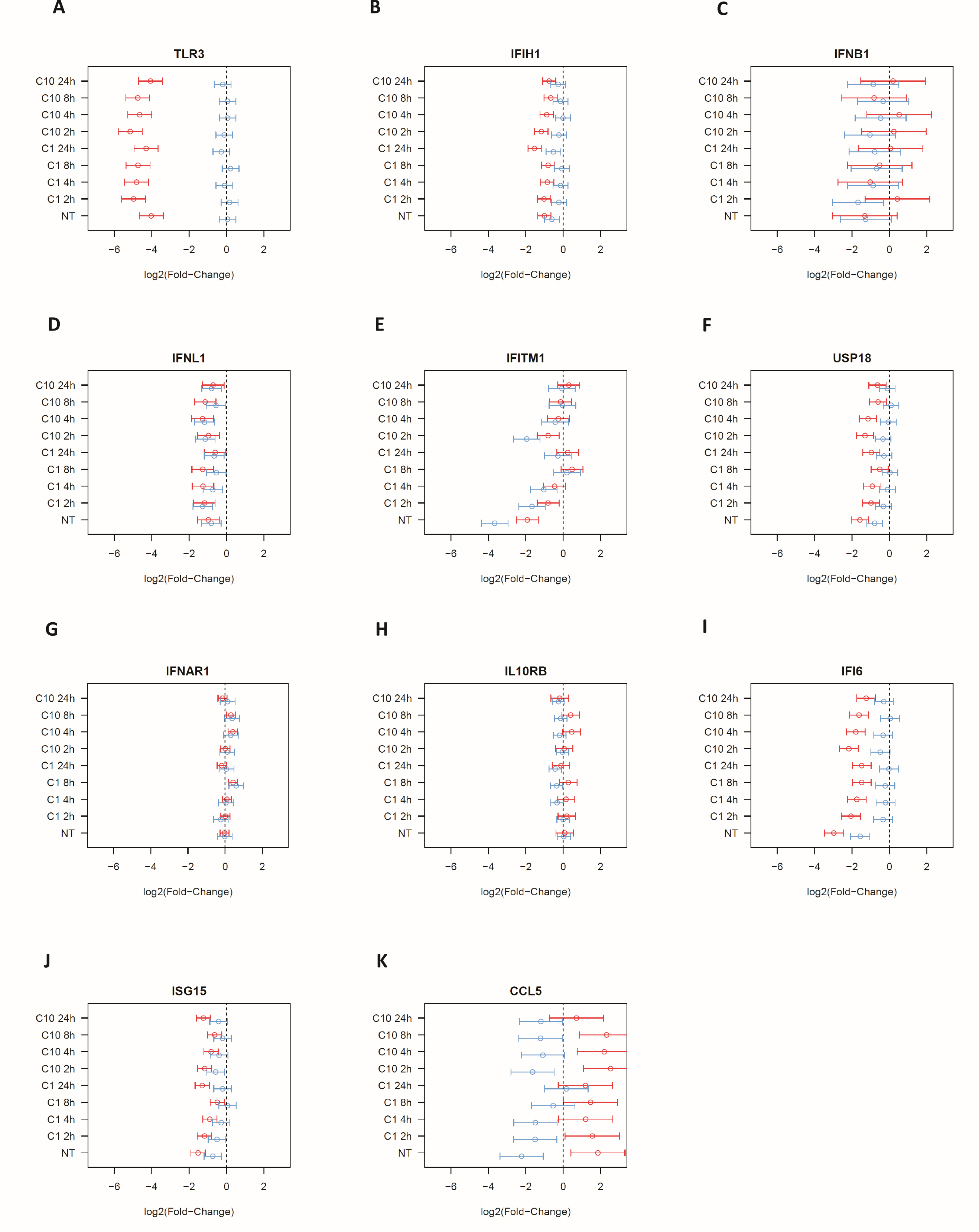

**Figure S7. Comparison of the impact of IRF6 and IRF8 on the activation of interferon responsive genes (ISGs) in IEC models.** Confidence Intervals relative to IFNβ treatments shown in **Fig. 3 and Fig. S6**. Confidence intervals for IRF6 and IRF8 are represented by blue and red bars, respectively. For raw data, see gene specific tables in **Supplementary Data Content 5**.

Alt text: This is a graphical representation of a comparison of the effects of IRF6 and IRF8, on the response to IFNβ treatments.
